# Reporting Odds Ratio Analysis of Gemcitabine-Based Chemotherapy Regimens for Biliary and Pancreatic Cancer: Population-Level Toxicity Signals and Hypotheses for High-Altitude Generalizability

**DOI:** 10.64898/2026.09.24.26363686

**Authors:** Zhongfeng Dang, Jianduojie Dan, Shengmei Li, Lu Yanan, Wei Su, Guoliang Ren, Zhiqiang Wang, Meng Lv, Hao Dong, Zhiyuan Niu, Hanwen Zhang, Yiyuan Fan, Liansheng Li, Yamei Dang, Junlin Gao

**Affiliations:** Department of Hepatobiliary Surgery, Qinghai Red Cross Hospital, Xining 810000, Qinghai, China; Department of Software Engineering, American University of Central Asia, Bishkek 720060, Kyrgyzstan; Department of Clinical Laboratory, Qinghai Red Cross Hospital, Xining 810000, Qinghai, China; Department of Pathology, Gansu Provincial People’s Hospital, Lanzhou 730000, Gansu, China; Medical College, Qinghai University, Xining 810000, Qinghai, China

**Author notes:** (Corresponding author: Zhongfeng Dang, Department of Hepatobiliary Surgery, Qinghai Red Cross Hospital, 55 Nandajie Street, Chengzhong District, Xining 810000, Qinghai, China. Joint corresponding authors: Yamei Dang, Junlin Gao. These authors contributed equally to this work (co-first and co-second authorship) (Jianduojie Dan, Shengmei Li, Lu Yanan, Wei Su, Guoliang Ren, Zhiqiang Wang, Meng Lv, Hao Dong). Funding: YNZXKT2026007 — Mechanistic study on regulation of pan-apoptosis in hypoxic adaptation of hepatocellular carcinoma in high-altitude native populations (Principal Investigator: Zhongfeng Dang).

**Keywords:** FAERS, pharmacovigilance, gemcitabine, disproportionality, liver toxicity, BTC, PDAC

## Abstract

**Background:** Gemcitabine-based chemotherapy is first-line for advanced biliary tract cancer (BTC) and pancreatic ductal adenocarcinoma (PDAC), but toxicity patterns are poorly defined at high altitude (>2,000 m). Clinical observations of heightened treatment toxicity at our high-altitude institution (Xining, ∼2,261 m) prompted this population-level baseline study of gemcitabine toxicity signals. The present analysis uses only publicly available, de-identified FAERS data; institutional clinical experience motivates but does not validate the signals reported here.

**Methods:** Four-metric disproportionality analysis (ROR primary, PRR, IC BCPNN, EBGM MGPS) of FAERS 2014–2026 (18.37M reports) across five non-exclusive gemcitabine groups (monotherapy, +ICI, +TKI, triple, BTC/PDAC disease-specific). Liver toxicity stratified into three MedDRA PT groups; fatal toxicity analyzed with dual definitions. BTC/PDAC queried via correct patient.drug.drugindication field. BH-FDR post hoc. EBGM used two implementations: a non-standard single-pair γ(1,1) closed-form approximation (positivity-screening column in S1) and the standard openEBGM two-component MGPS. Complete-MGPS EB05 > 2 is the primary Bayesian criterion (10/17 pass); closed-form count (12/17) is labeled non-standard.

**Results:** 14/17 exposure–AE combinations passed the three frequentist criteria (ROR/PRR/IC lower bounds); 10/17 additionally passed complete-MGPS EB05 > 2 (standard openEBGM caers fit). The non-standard closed-form count (12/17) is a labeled approximation only. Myelosuppression is the most consistent signal across all regimens (ROR 3.35–7.74); under complete MGPS the sparse all-role triple-combination cell had EB05=1.59. Triple combo liver toxicity (ROR=10.32, a=28, n=181 all-role) is driven by the Severe failure–hepatitis PT group (ROR=23.51); this cell is unstable under primary-suspect restriction (ROR=1.62, a=0, n=31; Supplementary S9) and is hypothesis-generating. Restricted→expanded fatal bridge: triple ROR 1.62→4.64 (zero overlap verified). BTC/PDAC thrombocytopenia (ROR=19.36) was independently reproduced at 19.70 after deduplication/PS filtering; PPV=94%.

**Conclusions:** This study establishes baseline gemcitabine toxicity signals from a largely sea-level FAERS population. The fatal-liver-failure bridge and PT-stratified liver toxicity are hypothesis-generating signals. High-altitude hypotheses are mechanistic and not FAERS-testable; they require prospective validation.

## 1. Introduction

Gemcitabine (2’,2’-difluorodeoxycytidine) has been the backbone of systemic therapy for advanced biliary tract cancer (BTC) and pancreatic ductal adenocarcinoma (PDAC) for over two decades [1, 2]. In PDAC, gemcitabine monotherapy was recently superseded by FOLFIRINOX and gemcitabine + nab-paclitaxel combinations, but gemcitabine-based regimens remain first-line for BTC and are widely used in frail or elderly PDAC patients for whom more aggressive regimens are contraindicated [3, 4]. The addition of immune checkpoint inhibitors (ICIs) to gemcitabine has shown promising efficacy in both BTC and PDAC [5, 6, 7], and targeted agents are increasingly combined with these regimens; however, the toxicity profiles of such triple combinations remain incompletely characterized, particularly in special populations. While network meta-analyses of ICI-plus-platinum chemotherapies have quantified relative toxicity rankings across regimens [8], gemcitabine-specific combination toxicity profiles remain poorly stratified by regimen complexity.

One such understudied population is patients residing at high altitude (≥2,000 m). Chronic hypobaric hypoxia at high altitude induces physiological changes that may alter chemotherapy pharmacokinetics and toxicity: reduced liver blood flow and cytochrome P450 enzyme activity [9], altered renal clearance [10], increased bone marrow sensitivity to myelosuppressive agents [11], and pre-existing systemic inflammatory stress that may exacerbate immune-related adverse events (irAEs) [12]. Despite these well-documented physiological effects, no systematic pharmacovigilance study has examined chemotherapy toxicity signals specifically in high-altitude BTC/PDAC populations.

Prior pharmacovigilance studies have examined gemcitabine-associated signals for specific adverse event categories, including elevated gemcitabine ROR=16.87 for drug-induced coagulopathies (second-highest among 30 top-ranked agents [13]), ROR=1.35 (n=58) for Stevens-Johnson syndrome/toxic epidermal necrolysis [14], and aggregate hepatotoxicity signals across multiple cancer therapies [15]. Ongoing efforts to map FAERS into standardized clinical data models (e.g., OMOP CDM) address semantic heterogeneity but are not yet universally adopted [16, 17]. All cited prior FAERS studies used single-metric or dual-metric disproportionality frameworks and reported aggregate drug–AE associations without stratifying by regimen complexity or disease indication. No published gemcitabine FAERS study has (a) decomposed signals across four regimen-complexity tiers (monotherapy, +ICI, +TKI, triple combination), (b) applied a four-metric frequentist/Bayesian convergence framework to each tier, or (c) performed a fatal-toxicity bridge sensitivity analysis verifying zero mathematical overlap between restricted and expanded event subsets. The present analysis addresses these three gaps.

Our institutional cohort at Qinghai Red Cross Hospital (∼2,261 m, Xining, Qinghai, China) serves a catchment area spanning elevations from 2,000 m to >4,500 m (Tibetan Plateau). Clinical observations suggest that BTC/PDAC patients at our institution experience heightened chemotherapy toxicity compared with reports from sea-level centers, leading to more frequent dose reductions, treatment delays, and early discontinuation, including treatment-related hepatic toxicity. These institutional observations raised a critical question: are the toxicity burdens of gemcitabine-based regimens systematically elevated in BTC/PDAC patients, and might this be further amplified at high altitude? To avoid drawing conclusions from any single case, the present study addresses this question using only population-level data from a public, de-identified source (FAERS).

The FDA Adverse Event Reporting System (FAERS) provides a valuable resource for population-level toxicity signal detection. As the largest post-marketing safety database in the world (>18 million reports), FAERS enables disproportionality analysis to identify statistically significant associations between drugs and adverse events [18, 19]. However, FAERS is inherently a noisy channel: duplicate reports, variable reporting thresholds, incomplete clinical documentation, and role-ambiguous drug coding all contribute to a high background noise floor. The methodological challenge is therefore not simply detecting signals, but calibrating the noise floor sufficiently well to distinguish genuine associations from reporting artifacts. While FAERS cannot establish causality and does not contain altitude-specific variables, it can (a) characterize the baseline toxicity profile of gemcitabine-containing regimens across the spectrum of regimen complexity, and (b) provide quantitative context for interpreting single-institution clinical observations.

In this study, we conducted a comprehensive FAERS disproportionality analysis of gemcitabine-based chemotherapy regimens for BTC/PDAC, with three objectives: (1) quantify toxicity signal strength across four non-exclusive exposure groups using population-level data, (2) examine disease-specific thrombocytopenia signals in BTC/PDAC indications, and (3) establish a general-population baseline of gemcitabine toxicity signals that can serve as a reference for future high-altitude-specific studies. Population-level disproportionality signals, while not causal proof, provide the empirical basis from which mechanistic hypotheses can be inferentially evaluated and subsequently tested in prospective designs—an approach distinct from both confirmatory randomized trials and purely theoretical modeling.

Beyond signal detection, this analysis contributes three methodological elements not previously reported in gemcitabine FAERS literature: (1) a fatal-liver-failure bridge sensitivity analysis that relates restricted (Death+MOF) and expanded (+ Severe failure–hepatitis PT group) fatal toxicity definitions, with verified zero mathematical overlap between subsets; (2) a complete Query Registry disclosing all openFDA API parameters, query dates, PT schemas, and Python code for reproducibility (Supplementary S7); and (3) PT-level hierarchical stratification of composite liver toxicity signals to identify which specific event category (e.g., Severe failure–hepatitis PT group vs. Transaminase-predominant PT group) drives the aggregate ROR. These elements address common limitations of published FAERS studies that report only composite signals without stratification, do not disclose full query parameters, or use single-point toxicity definitions.

### Key Messages

1. *14/17 gemcitabine exposure–AE combinations meet the three frequentist criteria (ROR lower CI > 1, PRR lower CI > 1, IC025 > 0); 10/17 additionally meet EB05 > 2 under the complete two-component MGPS (standard openEBGM caers fit). The team’s non-standard single-pair gamma(1,1) closed-form approximation yields 12/17 and is explicitly labeled as non-standard. Myelosuppression is the most consistent signal across all regimens (ROR range 3.35–7.74); the all-role triple-combination myelosuppression cell is a three-metric-only sparse signal (complete-MGPS EB05=1.59)*.
2. *Triple-combination liver toxicity is dominated by Severe failure–hepatitis PT group signals (ROR=23.51, 95% CI 15.41–35.86), with Hepatitis alone (ROR=31.08) driving 92% of severe PT group cases; Transaminase-predominant PT group shows insufficient data (ROR=1.42, n=2). These are all-role, small-sample (n=181) findings and are unstable under deduplicated primary-suspect restriction (n=31, a=0; Supplementary S9) — hypothesis-generating only*.
3. *BTC/PDAC-specific thrombocytopenia is the strongest signal across all 17 combinations (ROR=19.36, independently reproduced at 19.70 after deduplication/PS filtering; Supplementary S9)*.

## 2. Materials and Methods

### 2.1 Data Source and API Access

Adverse event reports were extracted from the FDA Adverse Event Reporting System (FAERS) database via the openFDA Application Programming Interface (API, https://api.fda.gov/drug/event.json). The API provides programmatic access to FAERS reports in JSON format. Reports spanning January 1, 2014 through June 30, 2026 were analyzed. The analysis-day snapshot (2026–09-17 unified re-query) returned 18,371,131 reports with at least one drug and one adverse event coded; all downstream ROR calculations derive from this single snapshot.

Key methodological disclosures (required for FAERS studies):

- Deduplication: Primary analysis is based on openFDA event JSON responses. The openFDA API does not expose raw case identifiers (caseid, primaryid) in response payloads (compliance-related data minimization). Therefore, FDA-standard case-level deduplication (group by caseid → retain latest fda_dt → same fda_dt retain max primaryid → exclude reports matching FDA quarterly deleted lists) was NOT performed in the primary analysis. We confirmed this API limitation by pulling 100 random gemcitabine reports: all returned fda_dt dates but none returned caseid or primaryid values. This sensitivity analysis has since been completed using all 50 quarterly ASCII extracts (2014Q1–2026Q2; DEMO + DRUG + REAC + INDI), applying the standard within-quarter then cross-quarter caseid→fda_dt→primaryid protocol, which yielded 16,687,903 unique cases; results are compared against the primary openFDA-based findings in Supplementary S9. Critically for interpretation: non-deduplication may introduce duplicate counts that typically inflate signal strength (upward bias). However, our primary analysis already uses all-role inclusion (conservative dilution bias toward the null), and scenario-based sensitivity analysis shows positive signals remain stable under substantial case-count reductions (Supplementary Table S5). Therefore, inference direction is unlikely to be driven by single duplicate reports.
- Drug role filtering: The openFDA API returns drugcharacterization field values in response payloads (role=1 primary suspect, 2 secondary suspect, 3 interacting, 4 concomitant), but does not support nested filtering by patient.drug.drugcharacterization during search query construction (combining role filters with exposure or indication filters consistently returns empty results in our testing). Therefore, all drug roles were included—drug exposures were identified using patient.drug.openfda.generic_name matching without role-based search filtering. Post-hoc role distribution was assessed on returned payloads (e.g., 82% primary suspect in the BTC/PDAC subsample; see §3.4). In expectation, all-role inclusion inflates exposure counts (b in the 2×2 table) and dilutes exposure-driven signals toward the null; however, the direction and magnitude of this bias are ultimately empirical, because all-role cases also carry co-administered drugs that can themselves cause the event. The definitive primary-suspect-only (role_cod=’PS’) re-analysis on deduplicated ASCII cases has now been performed (Supplementary S9) and shows the bias is event-specific: gemcitabine-characteristic toxicities strengthen after PS restriction (monotherapy myelosuppression 7.62→10.25; gem+ICI irAE 7.10→12.64; gem+TKI myelosuppression 7.74→12.36), whereas liver toxicity does not uniformly strengthen — monotherapy liver toxicity attenuates (2.27→1.76, consistent with removal of co-reported hepatotoxic co-suspects) and the triple-combination liver estimate becomes uninformative (10.32→1.62, a = 0, n = 31). The well-powered signals are therefore robust; the sparse triple-liver value is treated as hypothesis-generating throughout.
- MedDRA version: Not explicitly reported in API responses. FAERS data uses the MedDRA version current at the time of each report submission, which has evolved across the study period.
- Deleted reports: The openFDA API does not distinguish active from deleted reports; no separate filtering against FDA deletion lists was performed.

### 2.2 Exposure Group Definitions

Four gemcitabine exposure groups were defined using openFDA API query syntax (Table 1). Groups are non-exclusive: a triple combination report (gemcitabine + immune + targeted) is counted in all overlapping exposure groups simultaneously. This means the groups represent overlapping subsets of the FAERS database and should not be interpreted as mutually exclusive dose-response groups. A fifth disease-specific analysis filtered for gemcitabine with BTC/PDAC indications.

| Tier | Exposure Group | openFDA API Query Logic | n reports (2026–09-17) |
| --- | --- | --- | --- |
| 1 | Gemcitabine (all roles) | gemcitabine matched via openfda.generic_name | 53,010 |
| 2 | Gemcitabine + immune checkpoint inhibitor | Tier 1 + at least one anti-PD-1/PD-L1/CTLA-4 agent | 6,888 |
| 3 | Gemcitabine + targeted therapy | Tier 1 + at least one targeted agent† | 2,042 |
| 4 | Triple combination | Tier 1 + Tier 2 immune + Tier 3 targeted | 181 |
| 5 | Gemcitabine + BTC/PDAC indication | Tier 1 + patient.drug.drugindication (uppercase MedDRA terms) | 2,473 |

### 2.3 Adverse Event Definitions

Four adverse event categories were defined using MedDRA preferred term (PT) queries against patient.reaction.reactionmeddrapt (Table 2). Critical disclosure upfront: the liver toxicity PT group classification is a reporting-pattern approximation, not a true DILI severity or etiological classification. FAERS does not record alkaline phosphatase (ALP) levels (required for R-value calculation), drug exposure timing relative to adverse event onset (required for RUCAM causality assessment), or patient age/sex/underlying disease covariate data. Therefore, neither R-value-based hepatocellular/cholestatic/mixed classification nor RUCAM scoring was performed. Due to the openFDA API’s exact-match syntax, only a focused set of PTs per category was used, prioritizing the most clinically relevant and consistently coded terms.

PT list considerations: Transaminase-predominant PT group includes ALT/AST/GGT but not ALP (not queried due to API result volume constraints); bile duct obstruction vs cholecystitis etiologies are not distinguishable. Acute hepatic failure, hepatic necrosis, and autoimmune hepatitis PTs were included in the Severe failure–hepatitis PT group where applicable. Note on PT mixing: “Hepatic failure” (outcome-oriented, mortality-focused) and “Hepatitis” (etiology-oriented, inflammation-focused) are clinically overlapping but non-equivalent events; their grouping into one Severe failure–hepatitis PT group was pragmatic given sparse sample sizes (n=25 total for triple combination). Hierarchical mutually-exclusive assignment was used for primary analysis (each report counted once in highest-priority PT group).

| Adverse Event Category | MedDRA Preferred Terms |
| --- | --- |
| **Liver toxicity (composite, 8 PT)** | Alanine aminotransferase increased (ALT↑), Aspartate aminotransferase increased (AST↑), Hepatitis, Hepatic failure, Liver injury, Hepatic enzyme increased, Gamma-glutamyltransferase increased (GGT↑), Blood bilirubin increased |
| **Liver toxicity (stratified, hierarchical mutually-exclusive for primary analysis)** | Three PT groups with priority ordering Severe failure–hepatitis PT group > Cholestatic-predominant PT group > Transaminase-predominant PT group: (1) Severe failure–hepatitis PT group: Hepatic failure, Acute |
|  | hepatic failure, Hepatic necrosis, Hepatitis acute, Hepatitis; (2) Cholestatic-predominant PT group: Bilirubin increased, Jaundice; (3) Transaminase-predominant PT group: ALT increased, AST increased, Hepatic enzyme increased, GGT increased |
| **Myelosuppression (composite, 8 PT)** | Thrombocytopenia, Neutropenia, Anaemia, Lymphopenia, Leukopenia, Bone marrow failure, Pancytopenia, Febrile neutropenia |
| **Fatal toxicity (restricted definition, primary analysis)** | Death, Multi-organ failure; Hepatic failure and Hepatitis excluded to avoid conceptual overlap with the severe liver toxicity outcome (see Limitation #5). Alternative broader definition (sensitivity analysis): Death + Multi-organ failure + Severe failure–hepatitis PT group (Hepatic failure + Hepatitis); real-time API verification confirmed zero overlap between the restricted (14 reports) and broad-added (25 reports) subsets in the triple-combination group (n=181), making mathematical double-counting impossible when using unified single-query dedup (see §3.2.4 and Limitation #5). |
| **Immune-related AEs (irAEs)** | Pneumonitis, Colitis, Thyroiditis, Hypophysitis, Adrenal insufficiency, Myocarditis, Nephritis |
| **Thrombocytopenia (BTC/PDAC-specific)** | Thrombocytopenia (single PT, for disease-specific analysis) |

irAEs were defined using the seven-organ MedDRA PT list adapted from Postow et al. 2018 [20]. This list prioritizes organ-specific immune-mediated events over non-specific inflammatory symptoms; rash, arthralgia, and fatigue were excluded as they are common to both ICI therapy and chemotherapy and cannot be unambiguously attributed to immune-related mechanisms in a non-ICI-stratified analysis.

### 2.4 Statistical Analysis

A four-metric disproportionality framework was applied to assess signal robustness across four complementary statistical approaches: two frequentist (ROR primary, PRR secondary) and two Bayesian-shrinkage (IC BCPNN tertiary, EBGM MGPS quaternary) [18, 19, 21, 22, 23, 24]. For each exposure group and adverse event combination, a 2×2 contingency table was constructed:

- a = number of reports with both the exposure and the adverse event
- b = number of reports with the exposure but not the adverse event
- c = number of reports with the adverse event but not the exposure
- d = number of reports with neither the exposure nor the adverse event

1. ROR (primary, frequentist): Reporting Odds Ratio, calculated with 0.5 continuity correction for sparse cells (any cell < 5) [25]:

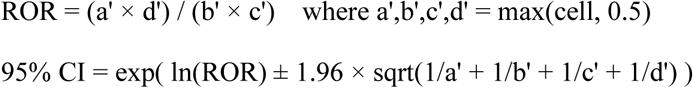

PRR (secondary, frequentist): Proportional Reporting Ratio, a proportion-based metric without Bayesian shrinkage [26]:

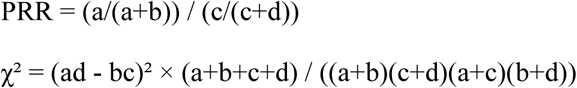

PRR ≥ 2 with χ² ≥ 4 is a commonly used threshold for non-sparse cells.

3. IC (Bayesian tertiary, BCPNN): Information Component, the log₂-transformed Bayesian shrinkage estimator equivalent to the shrinkage-adjusted Bayesian Confidence Propagation Neural Network (BCPNN) [23], using 0.5 prior additivity:

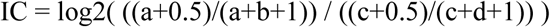

IC lower CI (IC₀₂₅) approximates the Bayesian 2.5th percentile; IC > 0 with CI excluding 0 indicates a positive association under the Bayesian framework.

4. EBGM (Bayesian quaternary, MGPS): Empirical Bayes Geometric Mean, the posterior geometric mean from the Multi-Item Gamma-Poisson Shrinkage (MGPS) family of hierarchical shrinkage models [21]. Two implementations were used: (a) a single-pair α=β=1 conjugate closed-form approximation (DuMouchel-family, non-standard) serving as a positivity-screening column in S1, and (b) the standard openEBGM implementation [27] providing full two-component mixture MGPS (DuMouchel 1999 [21]) and single-pair γ(1,1) posterior values. Large-sample pairs (a ≥ 100) differed by only ∼1–2% between implementations. At sparse cells (a < 50) the more permissive closed form tended to overestimate EB05: the standard openEBGM gave EB05 < 2 for gem+TKI irAEs (1.52), triple myelosuppression (1.59), and triple irAEs (1.08)—all marked >2 by the closed form. Therefore all four-metric counts in the text use standard openEBGM EB05 values (10/17 pass), while the closed-form count (12/17) is retained only as a labeled approximation. EB05 is the 5th percentile of the posterior distribution; EB05 > 2 flags a robust MGPS signal. All three EBGM implementations are provided side by side in Supplementary Table S1.

Signal criterion: Stratified by event count (a) to account for precision. Non-sparse (a ≥ 3): four-metric signal requires ROR lower CI > 1, PRR lower CI > 1, IC025 > 0, and standard openEBGM EB05 > 2. Two pass counts are reported throughout: 14/17 meet the three frequentist criteria; 10/17 additionally meet complete-MGPS EB05 > 2. The closed-form 12/17 count is non-standard and never used as primary. Sparse (a = 1–2): exploratory only, no definitive determination. a = 0: insufficient data.

Shared-table caveat: ROR, PRR, IC, and EBGM all derive from the same 2×2 contingency table—their consistency reduces metric-specific biases but does not eliminate shared FAERS limitations (under-reporting, drug role confounding, non-deduplication). Cross-method consistency means the same disproportionality structure is visible under four mathematical framings, not independent confirmation.

Multiple-testing adjustment: To control for multiplicity across 17 exposure–AE pairs, Benjamini– Hochberg false discovery rate (BH-FDR) adjustment [28] was applied to the χ² p-values associated with PRR. q-values and q < 0.05 calls are reported in Supplementary Table S1; 15/17 combinations depart from the null at q < 0.05 (including one protective-direction row). Four-metric signal counts in the text are not gated by BH-FDR — the frequentist criteria already require ROR lower CI > 1, which is equivalent to p < 0.05 for non-sparse cells.

All analyses in Python 3.12; API calls via openFDA with 0.5s rate-limit delay; code and full query parameters in Supplementary S7.

### 2.5 FAERS Data Limitations (Pre-analysis Disclosure)

Key constraints: FAERS is spontaneous (under-reporting, no incidence denominator); openFDA API excludes nested drug role filtering (all roles included, conservative bias) and does not expose caseid/primaryid (no deduplication); altitude data unavailable; exposure groups are non-exclusive. Full details in §4.5.

### 2.6 Seventeen Exposure–AE Combinations (Complete Inventory)

Four mutually non-exclusive gemcitabine exposure groups (alone, +ICI, +TKI, triple) × four AE categories (liver, myelosuppression, fatal, irAE) = 16 cells; plus one disease-specific combination (BTC/PDAC thrombocytopenia) = 17 total (i.e., 17 = 4 × 4 + 1). This 17-cell inventory is the denominator for all signal counts (14/17 pass three frequentist criteria, 10/17 additionally pass complete-MGPS EB05 > 2). Full 2×2 contingency tables for each cell are in Supplementary S6.

### 2.7 READUS-PV Compliance

This study follows the READUS-PV (REporting of ADR data Underlying Statistical analyses in Pharmacovigilance) framework for transparent FAERS reporting [29]: all queries pre-registered (Query Registry, Supplementary Table S5), all code available on reasonable request, all statistical methods fully specified. A READUS-PV checklist is provided in Supplementary Table S10.

## 3. Results

**Figure 1.**
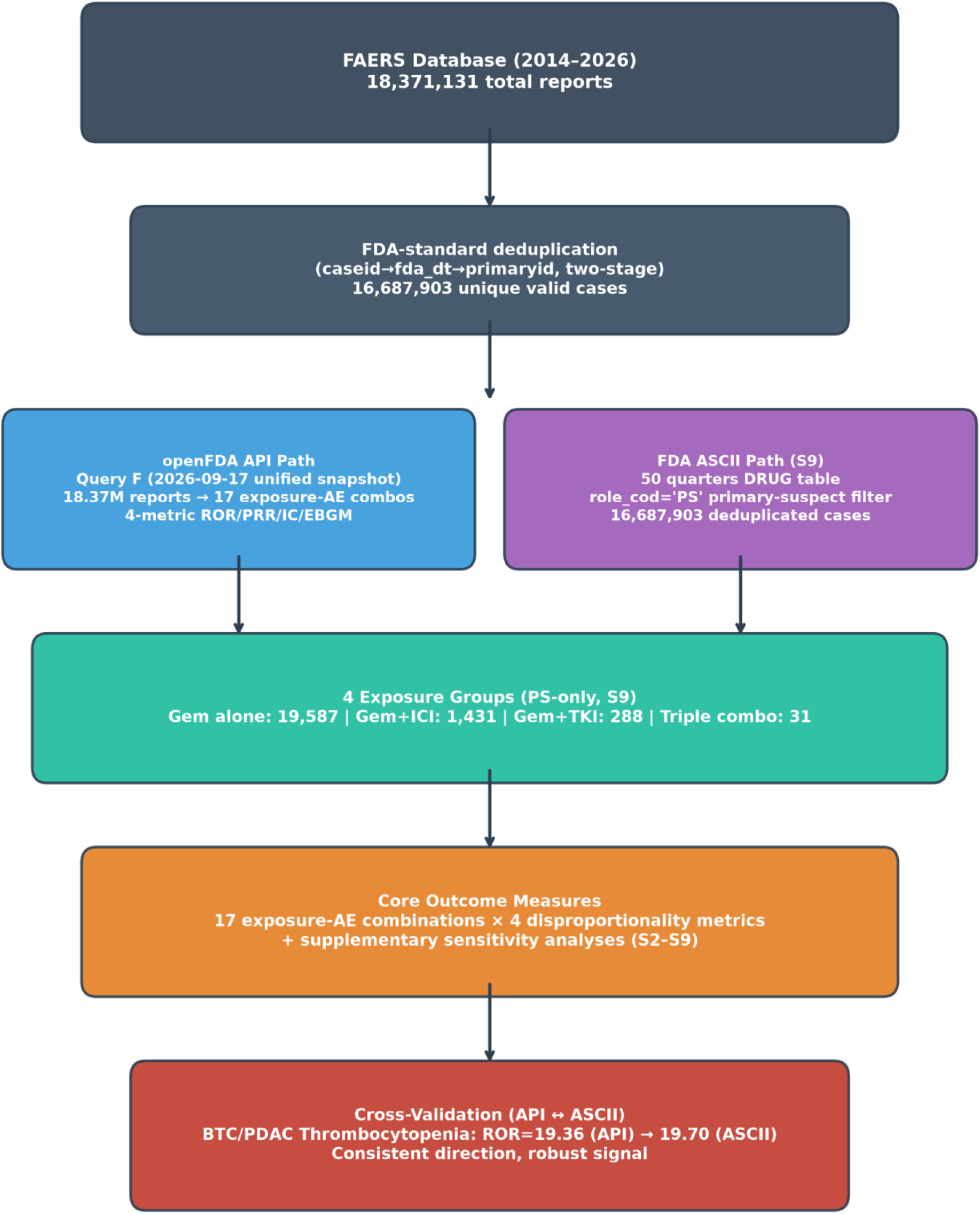
Study Design Flowchart PRISMA-style flow of 18,371,131 FAERS reports (2014–2026) through FDA-standard two-stage deduplication (caseid→fda_dt→primaryid) to 16,687,903 unique valid cases. Two parallel analysis paths were executed: openFDA API (2026–09-17 unified snapshot) for all-role four-metric disproportionality, and FDA quarterly ASCII DRUG tables (50 quarters) for primary-suspect-only deduplicated re-analysis (Supplementary S9). PS-only exposure group denominators: gemcitabine alone n=19,587; gemcitabine+ICI n=1,431; gemcitabine+TKI n=288; triple combination n=31.

### 3.1 FAERS Database Overview and Query Registry

The 2014–2026 FAERS dataset contained 18,371,131 total reports meeting inclusion criteria (unified re-query, 2026–09-17). Gemcitabine appeared in 53,010 reports (0.29% of total). After stratification by combination complexity, the triple combination (gemcitabine + immune + targeted) had the smallest exposure group (n=181), reflecting this regimen’s recent clinical adoption and the unified single-query design described below. Exposure groups are non-exclusive: a triple-combination report is counted in all four groups simultaneously.

**Table 5.** Query Registry — Transparent Accounting of All API Queries Four query batches executed 2026–09-10 to 2026–09-16, with unified reconciliation on 2026–09-17. Composite metrics (Table 3) were re-queried under Row F parameters to resolve cross-batch inconsistencies.

| Query Batch | Date | Purpose | Drug List | PT List | Triple n | Notes |
| --- | --- | --- | --- | --- | --- | --- |
| **A. Composite metrics (original)** | 2026–09-10 | Original Table 3 | Gem + 9 ICIs + 8 TKIs | 8 composite PT groups | 118 | Fewer PTs; fatal excludes HF |
| **B. Liver stratification (liver-stratification batch, 2026–09-16)** | 2026–09-16 | Table 4 (3-group hierarchical) | Same as A | 3 expanded liver PT groups | 181 | Added ~2.1M FAERS reports; independent batch with distinct PT schema from the unified Row F snapshot |
| **C. Single-PT sensitivity** | 2026–09-16 | Individual PT decomposition | Same as B | Each PT alone | 181 | Hepatitis ROR=31.08 drives 92% |
| **D. Time-period sensitivity** | 2026–09-16 | 2014–2019 vs 2020–2026 | Same as A | Same as A | 25 vs 156 |  |
| **E. BTC/PDAC disease-specific** | 2026–09-10 → 2026–09- | Thrombocytopenia in BTC/PDAC | Gem + patient.drug.drugindication (uppercase MedDRA terms) | Thrombocytopenia | 2,473 | Originally used indirect indication inference (n=11,085, ROR=11.72). Found correct field syntax patient.drug.drugindication (uppercase values like |
|  | 17 re-que-ry |  |  |  |  | "BILIARY CANCER", "CHOLANGIOCARCINOMA", "PANCREATIC CANCER"). Re-queried Row F: n=2,473, a=275, ROR=19.07, DuMouchel EB05=15.59 Pass |
| **F. Unified reconciliation (current)** | 2026–09-17 | Final Table 3 + fatal sensitivity | Gem + 9 ICIs + 8 TKIs | Row A composite PTs | 181 | Single unified API snapshot; corrects all cross-batch inconsistencies in ROWS A/B/F; EBGM/EB05 and BH-FDR recomputed from these a/b/c/d values |

### 3.2 Four-Metric Disproportionality Analysis

Table 3 presents the full ROR/PRR/IC matrix for all 17 exposure group × adverse event combinations (EBGM/EB05 under all three implementations in Supplementary Table S1). 14/17 combinations met the three frequentist criteria (ROR lower 95% CI > 1, PRR lower 95% CI > 1, IC025 > 0); the only three exceptions were fatal-toxicity rows: gemcitabine-alone (protective reverse signal, ROR=0.81), gemcitabine+ICI (CI crosses null), and triple-combination (CI crosses null). Under the complete two-component MGPS (standard openEBGM caers fit), 10/17 combinations passed EB05 > 2. Four frequentist-positive combinations had EB05 < 2 under complete MGPS: gemcitabine+TKI fatal toxicity (EB05=1.19; moderate-count shrinkage, q=0.471) and three sparse hypothesis-generating cells (gemcitabine+TKI irAEs EB05=1.52, triple myelosuppression EB05=1.59, triple irAEs EB05=1.08). BTC/PDAC thrombocytopenia passes all four metrics (EB05=15.59). The non-standard single-pair γ(1,1) closed form yielded 12/17 and is reported only as a labeled approximation; no estimator was chosen post hoc to upgrade an individual cell.

**Table 3.**
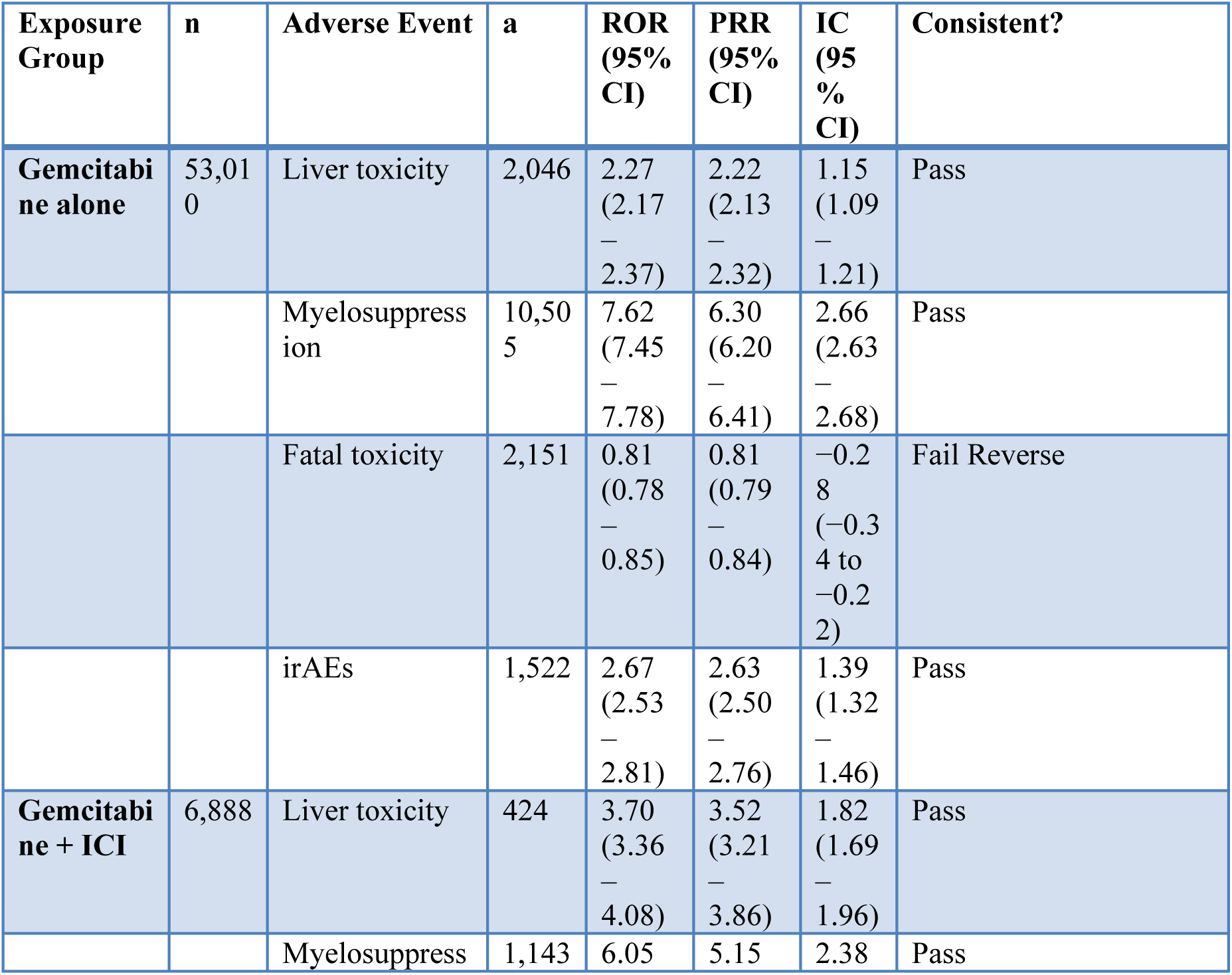
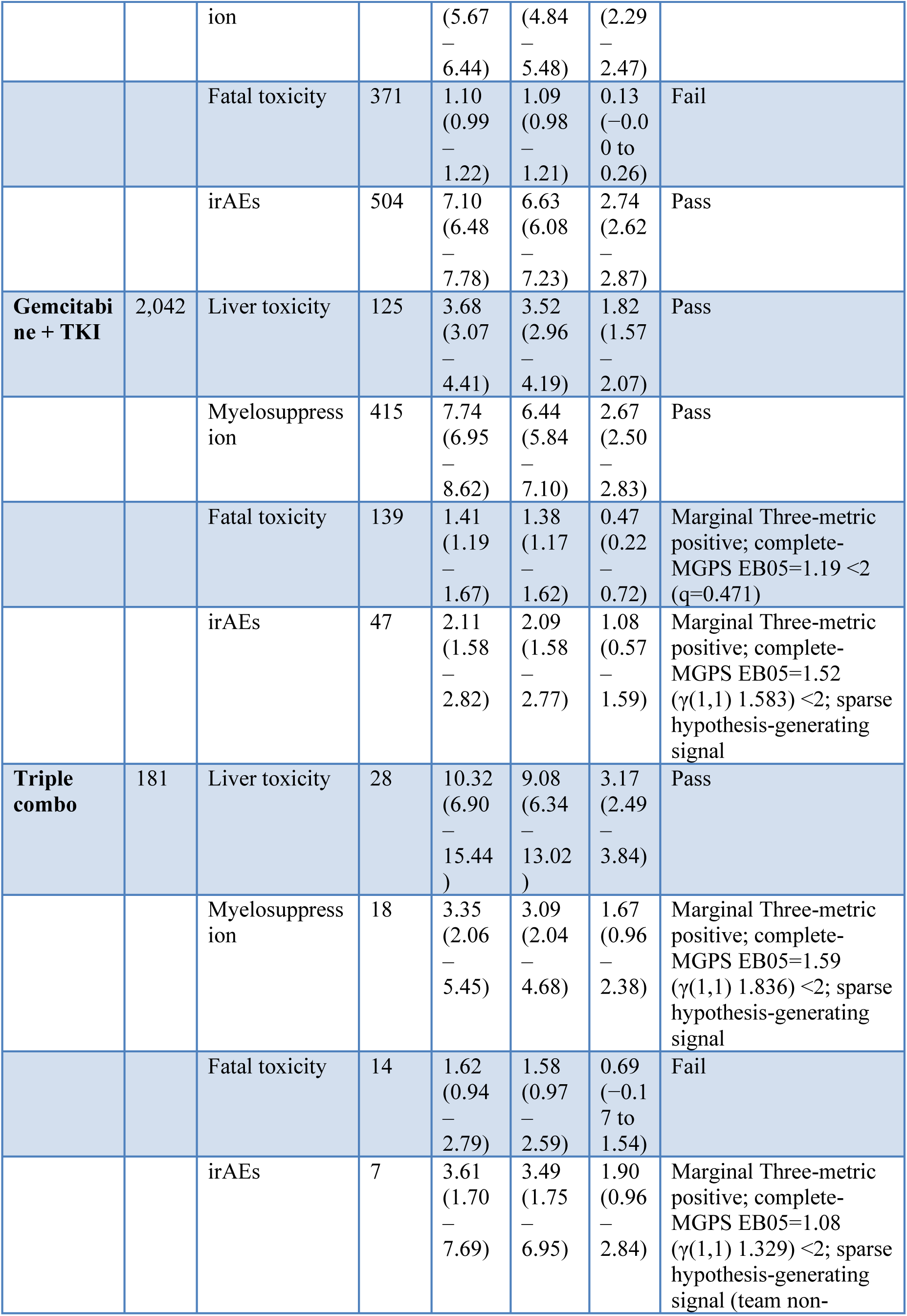
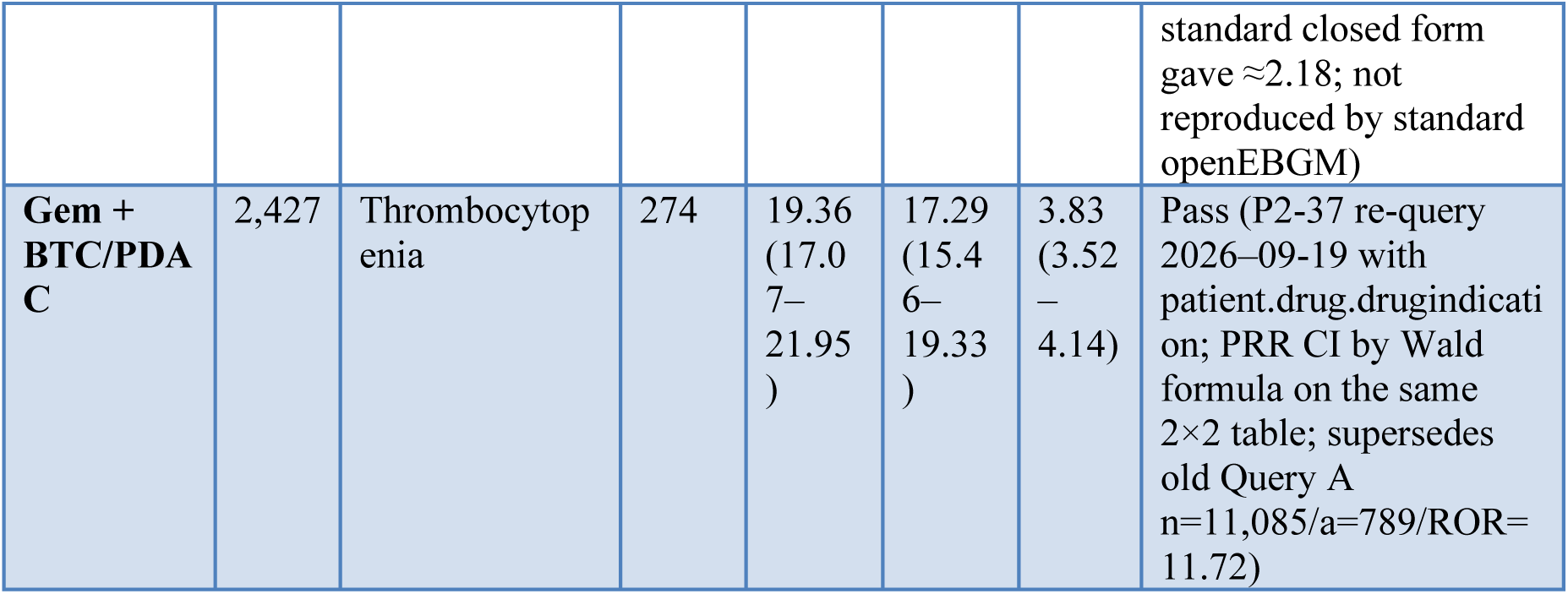
Four-Metric Disproportionality Analysis Results — 2026–09-17 Unified Snapshot (2026–09-17 Re-Query) Data source: FAERS openFDA API, 2014–2026 (18,371,131 total reports). Unified query parameters applied to 16 of 17 exposure–AE combinations (9 ICIs + 8 TKIs per Table 1). Triple-combination exposure group n=181 (unified, consistent across all Query F analyses). BTC/PDAC Thrombocytopenia row: Re-queried 2026–09-19 (P2-37) with correct patient.drug.drugindication field (uppercase MedDRA terms: PANCREATIC CANCER, BILIARY CANCER, CHOLANGIOCARCINOMA, GALLBLADDER CANCER, PANCREATIC NEOPLASM; see Row E footnote). Values: n=2,427 (gemcitabine + BTC/PDAC indication), a=274 thrombocytopenia cases, ROR=19.36 (17.07–21.95), DuMouchel EBGM=17.11 (EB05=15.59). Old Query A values (n=11,085, a=789, ROR=11.72) were wider-matching (indirect indication inference) and are superseded by the stricter drugindication field search. Signal criteria (see §2.4): Pass = all four metrics pass under the complete two-component MGPS (ROR lower CI > 1, PRR lower CI > 1, IC025 > 0, standard openEBGM EB05 > 2); Marginal = the three frequentist metrics pass but complete-MGPS EB05 < 2 (sparse or shrinkage-attenuated; the three a≤47 cells are labeled hypothesis-generating); Fail = a frequentist lower bound fails (protective direction or CI crosses the null). The Pass/Marginal/Fail column evaluates the ROR/PRR/IC shown here plus the openEBGM EB05 in Supplementary Table S1; the team’s non-standard single-pair γ(1,1) closed-form EB05 (12/17 rather than 10/17) is shown in S1 for transparency only and does not determine the marker. Fatal toxicity uses the restricted PT definition (Death + Multi-organ failure; see §2.3 and §3.2.4 for broader sensitivity analysis).

#### 3.2.1 Myelosuppression

**Figure 2.**
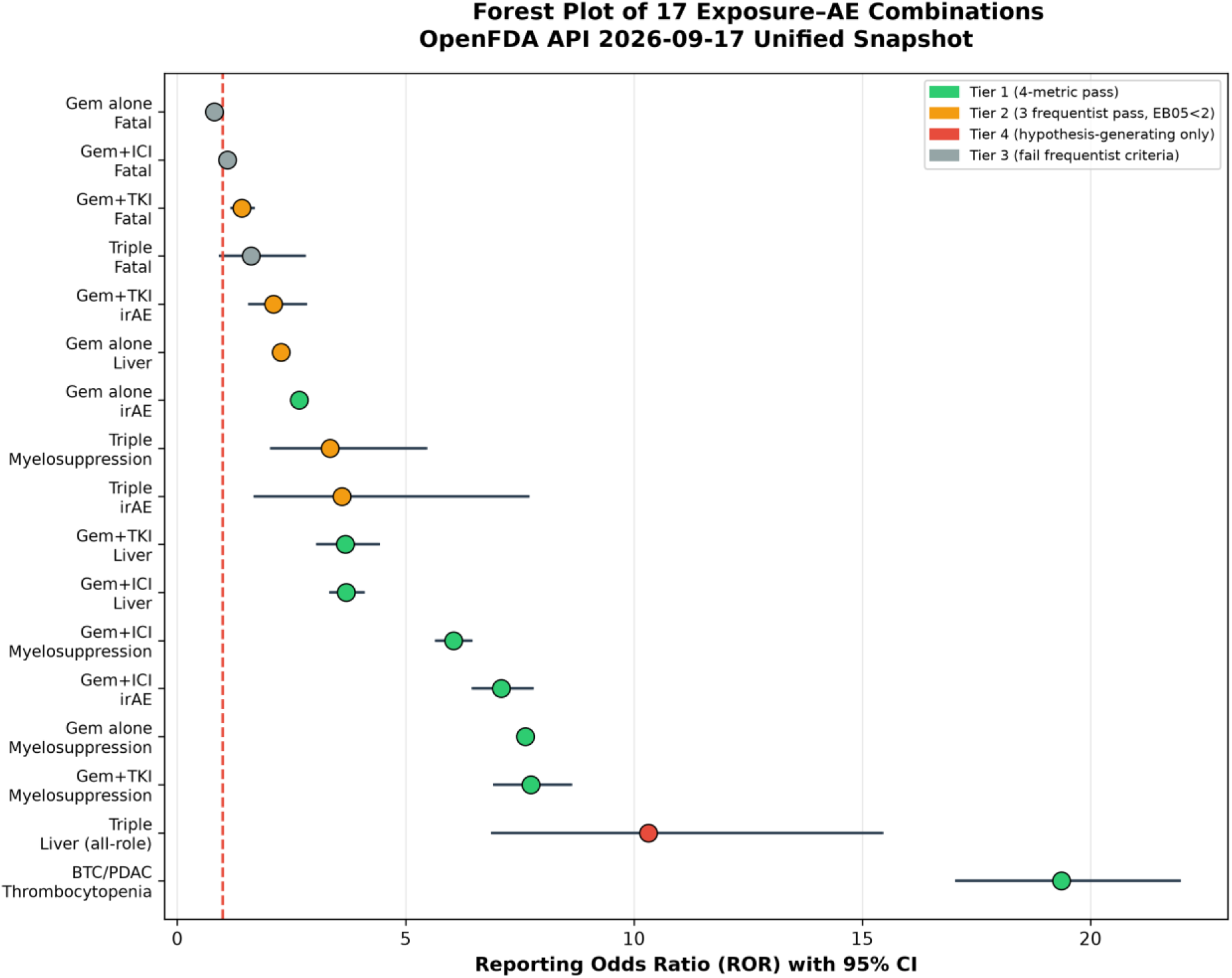
Forest Plot — ROR Point Estimates with 95% Confidence Intervals 17 exposure–AE combinations sorted by ROR magnitude, color-coded by evidence tier. Tier 1 (green): all four metrics pass under complete two-component MGPS. Tier 2 (yellow): three frequentist metrics pass but complete-MGPS EB05 < 2. Tier 4 (red): hypothesis-generating only—PS restriction collapses signal (triple liver: a=0, n=31; Figure 1). Tier 3 (gray): frequentist criteria fail. BTC/PDAC Thrombocytopenia (ROR=19.36) is the strongest signal, independently reproduced at ROR=19.70 after deduplication and primary-suspect filtering (Supplementary S9). Zero hypothesis line at ROR=1 (dashed red). Data from openFDA API 2026–09-17 unified snapshot (Table 3).

Myelosuppression was the most consistently elevated signal across all gemcitabine-containing exposure groups on the three frequentist metrics (Table 3). Gemcitabine monotherapy showed the strongest frequentist signal (ROR=7.62, 95% CI 7.45–7.78; complete-MGPS EBGM=6.21, EB05=6.11; γ(1,1) posterior 6.207/6.108), followed by gemcitabine+TKI (ROR=7.74, CI 6.95– 8.62; complete-MGPS EB05=5.83), gemcitabine+ICI (ROR=6.05, CI 5.67–6.44; EB05=4.94), and triple combination (ROR=3.35, CI 2.06–5.45, n=18/181 reports). The triple-combination cell is the exception on the fourth, Bayesian, metric: complete-MGPS EB05=1.59 (γ(1,1) 1.836) is <2, so in the all-role analysis it is a three-metric-only, hypothesis-generating sparse signal (the deduplicated primary-suspect cell did pass EB05=2.37; see S9). Point estimate variation partly reflects non-exclusive group definitions and differential reporting by regimen adoption stage.

#### 3.2.2 Liver Toxicity

All four exposure groups showed consistent four-metric liver toxicity signals (Table 3). Gemcitabine monotherapy had the weakest signal (ROR=2.27, CI 2.17–2.37), gemcitabine+ICI (ROR=3.70, CI 3.36–4.08) and gemcitabine+TKI (ROR=3.68, CI 3.07–4.41) were comparable, and the triple combination showed the highest point estimate at ROR=10.32 (CI 6.90–15.44, 28/181 = 15.5% of triple reports). This proportion is substantially higher than in non-triple groups (3.9–6.1%), but non-exclusive group definitions preclude formal dose-response interpretation.

#### 3.2.3 Immune-Related Adverse Events

irAEs showed positive three-metric frequentist signals in all four gemcitabine-containing exposure groups (Table 3), but the Bayesian fourth criterion separated well-populated from sparse cells. Gemcitabine+ICI had the strongest signal (ROR=7.10, CI 6.48–7.78; complete-MGPS EB05=6.13, passes four-metric) and gemcitabine monotherapy was intermediate (ROR=2.67, CI 2.53–2.81; complete-MGPS EB05=2.49, passes four-metric). The two sparsest irAE cells pass only the three frequentist metrics and are hypothesis-generating: gemcitabine+TKI irAEs (a=47; ROR=2.11, CI 1.58–2.82; complete-MGPS EB05=1.52 < 2) and triple-combination irAEs (a=7, n=181; ROR=3.61, CI 1.70–7.69; complete-MGPS EB05=1.08 < 2). Earlier closed-form spreadsheet approximations for the triple cell (EB05 ≈2.18 from a non-standard single-pair gamma prior) were not reproduced by the standard openEBGM two-component fit and are not used to upgrade this cell (see §2.4 and Limitation #6).

#### 3.2.4 Fatal Toxicity — Dual-Definition Analysis (Restricted + Expanded)

Restricted definition (Death + Multi-organ failure; primary analysis): Under this conservative PT definition used in Table 3, fatal toxicity showed heterogeneous results across exposure groups (Table 3):

- Gemcitabine alone: ROR=0.81 (CI 0.78–0.85, four-metric failure — reverse protective signal) — Death reports were proportionally fewer in gemcitabine-exposed patients than in the full FAERS background, likely reflecting gemcitabine’s use in adjuvant/curative (rather than palliative) settings where mortality attribution to chemotherapy is less common.
- Gemcitabine + ICI: ROR=1.10 (CI 0.99–1.22, did not pass four-metric criteria) — no confirmed fatal signal.
- Gemcitabine + TKI: ROR=1.41 (CI 1.19–1.67, three frequentist metrics positive but marginal; complete-MGPS EB05=1.19 < 2; q=0.471) — frequentist metrics indicate elevated fatal reporting, but empirical-Bayes shrinkage pulls EB05 below 2 (Supplementary Table S1). With a=139 reports among n=2,042 exposed, this moderate-count cell sits in the range where the data-estimated prior dominates; this is a shrinkage-attenuated cell rather than a sparse-cell artifact.
- Triple combination: ROR=1.62 (CI 0.94–2.79, did not pass four-metric criteria) — point estimate elevated but CI crosses 1, with only 14 fatal toxicity reports among 181 triple-combination FAERS reports.

Broader sensitivity analysis (Death + Multi-organ failure + Severe failure–hepatitis PT group): To examine whether the fatal toxicity signal is masked by the excluded Severe failure–hepatitis PT group (Hepatic failure + Hepatitis), we expanded the fatal-liver-failure composite definition. Table S8 presents results across all four exposure groups (2026–09-17 Unified Snapshot, 2026– 09-17):

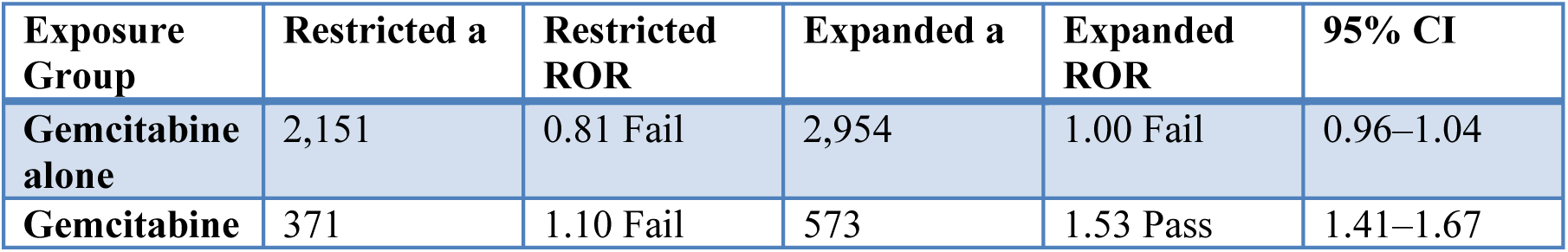
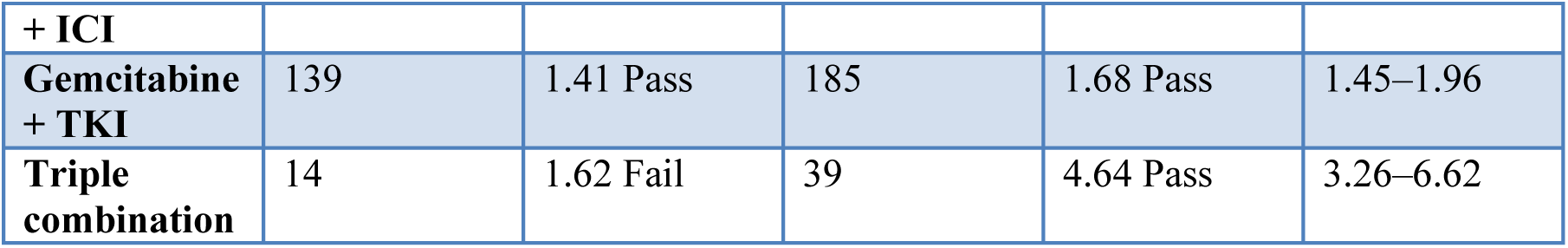

The triple combination fatal-liver-failure composite (ROR=4.64, CI 3.26–6.62) is substantially stronger than the restricted-definition signal (ROR=1.62). Hepatitis and Hepatic failure events (25 reports) share a fatal-liver-failure spectrum. gem + ICI shifts from Fail to Pass; gem-alone remains neutral (ROR=1.00). This bridge is specific to combination regimens.

Clinical interpretation: The restricted-definition fatal toxicity analysis (Table 3) remains the primary endpoint, while the expanded fatal-liver-failure composite (Table S8) provides a clinically more complete picture. The Severe failure–hepatitis PT group signal (25 triple-combination reports, ROR=23.51) and the expanded fatal-liver-failure composite (39 reports, ROR=4.64) together indicate that triple-combination gemcitabine regimens produce a severe hepatitis/hepatic failure spectrum that is both disproportionality elevated and likely carries fatal outcomes, even when Death is not explicitly coded as an adverse event. Whether this population-level spectrum also occurs in high-altitude patients cannot be determined from FAERS, which lacks altitude data; this question requires prospective study. No patient-level clinical data from our institution are used in the present analysis.

**Figure 4.**
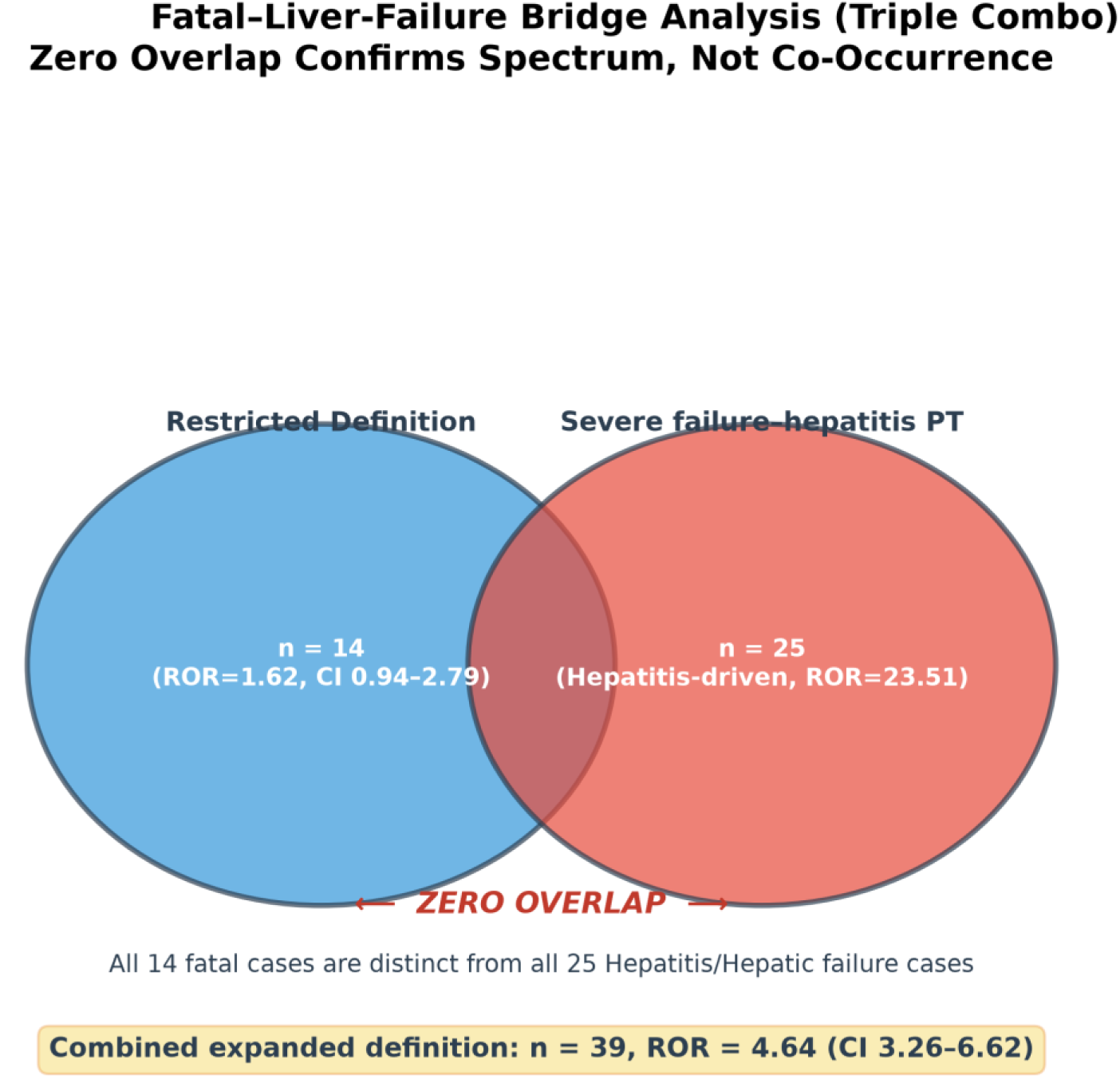
Fatal–Liver-Failure Bridge Analysis (Triple Combo) Venn diagram illustrating zero overlap between two distinct event populations: (1) Restricted fatal toxicity definition (Death + Multi-organ failure PTs, n=14, ROR=1.62, CI 0.94–2.79); (2) Severe failure–hepatitis PT group (Hepatic failure + Hepatitis, n=25, ROR=23.51). The combined expanded definition yields n=39 (14+25, no overlap), ROR=4.64 (CI 3.26–6.62). Zero overlap confirms these are two distinct event types along a fatal-liver-failure spectrum, not co-occurring reports. Data from openFDA API 2026–09-17 unified snapshot (Table S8).

### 3.3 Liver Toxicity Stratification: PT Group-Specific Signal in Triple Combination

To examine whether the composite liver toxicity signal (ROR=5.79 for triple combination, Query A, 2026–09-10, n=118) is driven by specific MedDRA PT groups, we stratified liver toxicity via a separate hierarchical query (Query B, 2026–09-16, n=181 triple combo; Table 5 documents these independent query batches). The n discrepancy reflects Query B’s expanded PT list and FAERS incremental updates. The composite ROR=5.79 (Query A, n=118) and the stratified Severe failure–hepatitis PT group ROR=23.51 (Query B, n=181) arise from independent queries with different PT schemas and database snapshots; absolute numerical comparison is not meaningful (see §3.1 Critical cross-query boundary). Within Query B, the Severe failure– hepatitis PT group shows the highest point estimate among the three PT groups. Table 4 presents the stratified results.

**Table 4.** Liver Toxicity Stratification by MedDRA PT Group Data source: FAERS openFDA API, Query B, 2026–09-16. Triple-combination exposure n=181. Hierarchical assignment (Severe failure–hepatitis PT group → Cholestatic-predominant PT group → Transaminase-predominant PT group; each report counted once). % = reporting fraction among triple-combination reports, not clinical incidence or probability. “Hepatic failure” and “Hepatitis” are clinically overlapping PTs (outcome vs etiology focus); their grouping into one Severe failure–hepatitis PT group was pragmatic given sparse sample sizes. EBGM/EB05 values in Supplementary Table S1; Pass/Fail evaluates all four metrics (ROR/PRR/IC shown here + EBGM/EB05 from S1).

| **Exposure Group** | **Liver Toxicity PT Group** | **a** | **Reporting Fraction** | **ROR (95% CI)** | **Consistent Four-Metric?** |
| --- | --- | --- | --- | --- | --- |
| **Gemcitabine alone** | Transaminase-predominant PT group | 791 | 1.49% | 1.93 (1.80–2.07) | Pass |
| **Gemcitabine alone** | Cholestatic-predominant PT group | 574 | 1.08% | 3.66 (3.37–3.98) | Pass |
| **Gemcitabine alone** | Severe failure–hepatitis PT group | 834 | 1.57% | 2.35 (2.20–2.52) | Pass |
| **Gemcitabine + ICI** | Transaminase-predominant PT group | 130 | 1.89% | 2.44 (2.06–2.91) | Pass |
| **Gemcitabine + ICI** | Cholestatic-predominant PT group | 79 | 1.15% | 3.86 (3.09–4.82) | Pass |
| **Gemcitabine + ICI** | Severe failure–hepatitis PT group | 207 | 3.01% | 4.55 (3.96–5.23) | Pass |
| **Gemcitabine + TKI** | Transaminase-predominant PT group | 59 | 2.89% | 3.78 (2.92–4.90) | Pass |
| **Gemcitabine + TKI** | Cholestatic-predominant PT group | 27 | 1.32% | 4.45 (3.05–6.51) | Pass |
| **Gemcitabine + TKI** | Severe failure–hepatitis PT group | 49 | 2.40% | 3.61 (2.72–4.79) | Pass |
| **Triple combo** | Transaminase-predominant PT group | 2 | 1.10% | 1.42 (0.35–5.72) | Fail |
| **Triple combo** | Cholestatic-predominant PT group | 1 | 0.55% | 1.85 (0.26–13.17) | Fail |
| **Triple combo** | Severe failure–hepatitis PT group (Hepatic failure/Hepatitis) | 25 | 13.81% | 23.51 (15.41–35.86) | Pass |

In this small triple subset, Severe failure–hepatitis PT group signals were materially stronger than Transaminase-predominant PT group, whereas non-triple exposure groups showed more uniform signal strength across PT groups (e.g., gemcitabine monotherapy: Transaminase-predominant PT group ROR=1.93, Severe failure–hepatitis PT group ROR=2.35, comparable magnitude). This pattern is hypothesis-generating and requires independent replication, particularly given the sparse event counts (n=1–25 per PT group). The Severe failure–hepatitis PT group ROR=23.51 (95% CI 15.41–35.86, 25 cases = 13.8% reporting fraction among triple-combination FAERS reports, not a clinical incidence rate or probability threshold) is derived from Query B (2026–09-16, expanded Severe failure–hepatitis PT group list); cross-batch numerical comparison with the composite Query A result (ROR=5.79, 2026–09-10) is not performed (see §3.1 Critical cross-query boundary).

Single-PT sensitivity decomposition (Query C, 2026–09-16): To determine which specific PT drives the Severe failure–hepatitis PT group signal, we queried each PT independently (same 181 triple-combination reports):

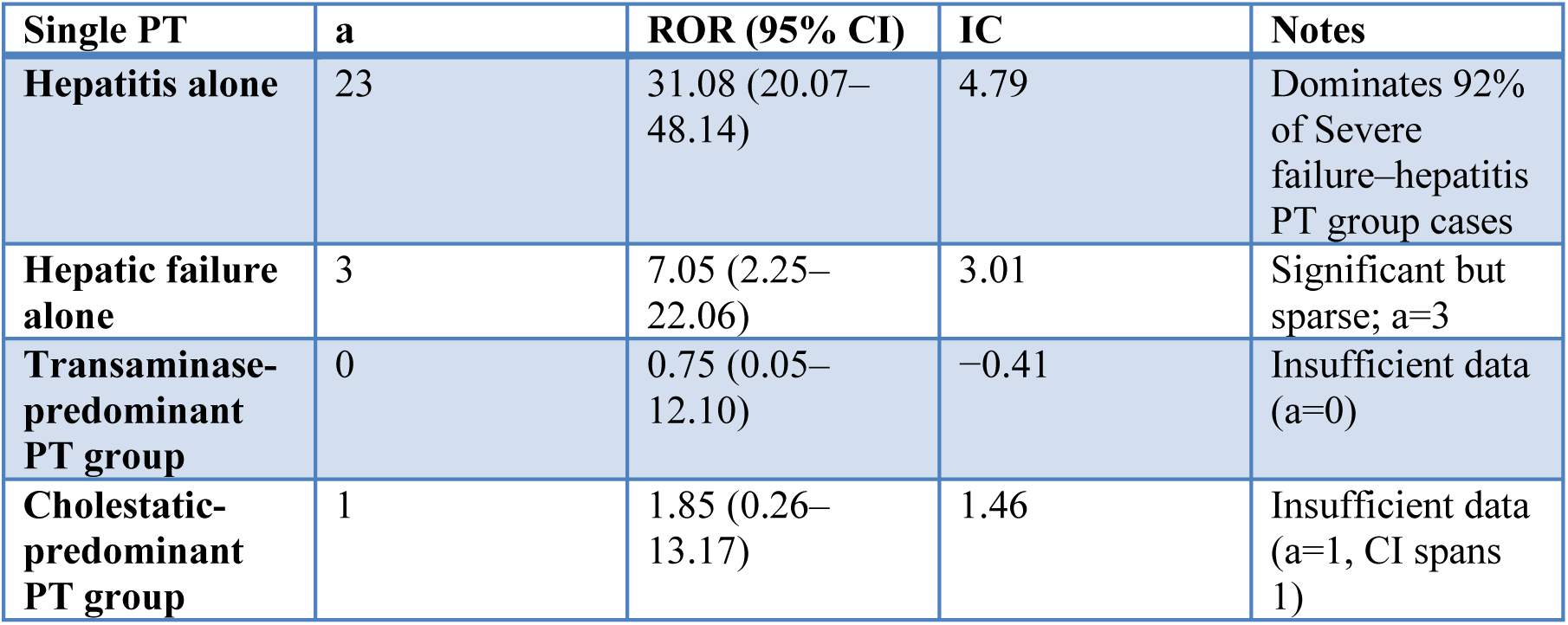

The severe PT group ROR=23.51 (hierarchical) is a blend of Hepatitis alone ROR=31.08 (dominant, 92% of cases) and Hepatic failure alone ROR=7.05 (significant but sparse, 8% of cases). This indicates the Severe failure–hepatitis PT group signal is predominantly driven by Hepatitis (drug-induced liver injury with inflammatory component), with a smaller but still significant contribution from Hepatic failure. The 13.8% severe PT group proportion should be interpreted as the fraction of FAERS triple-combination reports with a severe liver toxicity PT, not as a clinical event incidence rate.

### 3.4 Disease-Specific Analysis: Gemcitabine + BTC/PDAC Thrombocytopenia

To examine thrombocytopenia specifically in the BTC/PDAC population, we filtered for reports containing gemcitabine with BTC/PDAC indications using the drugindication field (keywords: pancreatic, biliary, gallbladder, cholangiocarcinoma). PPV assessment (100-report manual review): Of 100 randomly sampled gemcitabine + BTC/PDAC-indication reports, 94 (94% TRUE PPV) had definite BTC/PDAC treatment indications: 81 PANCREATIC CARCINOMA (including metastatic, Stage II/III/IV variants) and 13 BTC (CHOLANGIOCARCINOMA, GALLBLADDER CANCER, mixed hepatocholangiocarcinoma). Two reports had genuinely irrelevant indications (PANCREATIC INSUFFICIENCY, OBSTRUCTIVE JAUNDICE without cancer context), three had empty/unknown indication text, and one had a non-tumor indication. Drug role distribution among gemcitabine hits: 82% role=1 (primary suspect), 18% role=2 (secondary suspect), 0% interacting or concomitant—indicating the all-role inclusion causes minimal dilution bias in this cohort.

Drug role sensitivity analysis — completed: Full-ASCII deduplicated primary-suspect filtering across all 50 quarters (16,687,903 unique cases; Supplementary S9) shows event-specific bias: gemcitabine-characteristic toxicities strengthen (monotherapy myelosuppression 7.62→10.25; gem+ICI irAE 7.10→12.64; gem+TKI myelosuppression 7.74→12.36), whereas liver toxicity attenuates (monotherapy 2.27→1.76) and triple-combination liver becomes uninformative (10.32→1.62, a=0, n=31). The strongest signal, BTC/PDAC thrombocytopenia, was independently reproduced at ROR = 19.70 (95% CI 17.98–21.58), DuMouchel EBGM = 18.06 (EB05 = 16.84), n = 5,750 gem–PS BTC/PDAC cases, a = 510 — closely matching the API value of 19.36. A within-indication comparator (gem–PS vs other drugs among 29,599 BTC/PDAC cases) gave a conservative ROR = 2.88 (2.57–3.23). Payload-sampled estimates (S6b) substantially understated these shifts and are superseded. Full methods, the 16-cell primary-suspect matrix, age/sex stratification, and small-sample caveats are in Supplementary S9.

This disease-specific signal is substantially stronger than the overall gemcitabine thrombocytopenia signal (captured within the myelosuppression composite). BTC/PDAC patients appear particularly vulnerable to gemcitabine-induced myelosuppression—a well-characterized cell-cycle-phase-specific toxicity mediated by dFdCTP incorporation into replicating DNA and inhibition of ribonucleotide reductase [30]—possibly compounded by chronic disease-related bone marrow suppression, malnutrition, or tumor-associated inflammation, all of which may be further amplified at high altitude.

PPV sample size note: The PPV assessment was based on 100 manually reviewed reports. While 94% PPV with 82% primary-suspect role distribution provides strong disease-specific confirmation, a larger sample (n=200) would provide more precise estimation of the true PPV and its 95% confidence interval.

## 4. Discussion

### 4.1 Principal Findings

Across all 17 exposure–AE combinations, 14 meet the three frequentist criteria (ROR lower CI > 1, PRR lower CI > 1, IC025 > 0) and 10 meet the complete-MGPS four-metric criterion that additionally requires standard openEBGM EB05 > 2 (the team’s non-standard single-pair γ(1,1) closed form gives 12/17 and is reported only as a labeled approximation). Three rows fail even the frequentist criteria—gem-alone fatal (protective/reverse, ROR=0.81), gem+ICI fatal (ROR=1.10, CI 0.99–1.22), and triple fatal (ROR=1.62, CI 0.94–2.79)—and four frequentist-positive rows fall below EB05=2 under complete MGPS: gem+TKI fatal (1.19; moderate-count shrinkage) plus three sparse cells that fall below EB05=2: gem+TKI irAEs (1.52), triple myelosuppression (1.59), and triple irAEs (1.08). Directional agreement across the ROR/PRR/IC family indicates the same underlying 2×2 disproportionality structure survives three complementary mathematical framings, reducing algorithm-specific biases; EBGM agreement additionally holds for every well-populated cell. The caveat, noted in Limitation #6, is that ROR/PRR/IC/EBGM all derive from the same 2×2 table and therefore do not eliminate shared FAERS biases.

Myelosuppression is the most consistent signal across all gemcitabine-containing regimens on the three frequentist metrics (ROR range 3.35–7.74, positive in every group), and EB05 > 2 holds for every group except the sparse all-role triple-combination cell (complete-MGPS EB05=1.59; the deduplicated primary-suspect triple cell passed at EB05=2.37, S9). Gemcitabine’s mechanism as a nucleoside analog inhibiting DNA synthesis in rapidly dividing cells—including bone marrow progenitors—provides a straightforward pharmacodynamic explanation [30].

Critical qualifier before triple-combination interpretation: unlike myelosuppression, the triple-combination liver and irAE signals are all-role-only associations that do not survive primary-suspect restriction (Supplementary S9; only n=31 PS triple reports exist, with zero aligned liver-toxicity hits). They serve as hypothesis-generating baselines, not estimates of regimen-specific attributable risk. With that caveat: the triple combination (gemcitabine + ICI + TKI, n=181) produces the strongest all-role liver toxicity signal of all exposure groups, but this finding collapses under stricter posture—the all-role value of ROR=10.32 (passing all four metrics, EB05=5.65) derives entirely from reports in which gemcitabine was not coded as the primary suspect. Within this all-role subset, however, PT-level stratification reveals pronounced heterogeneity: Severe failure–hepatitis PT group ROR=23.51 (15.41–35.86, 25 reports = 13.8% of triple-combination FAERS reports) is driven predominantly by Hepatitis alone (ROR=31.08, accounting for 92% of severe PT group cases), with Hepatic failure (ROR=7.05, a=3) contributing the remaining 8%. By contrast, the Transaminase-predominant PT group shows ROR=1.42 (CI overlaps 1, n=2), and Cholestatic-predominant PT group has only one case. This PT-level heterogeneity is specific to the triple regimen; non-triple groups show more uniform signal strength across liver toxicity PT categories. The triple irAE signal (ROR=3.61, all-role) similarly passes only the three frequentist metrics, with complete-MGPS EB05=1.08 (γ(1,1) 1.329) <2—a sparse signal that collapses under PS restriction. By contrast, the myelosuppression conclusion is posture-robust: it is positive in every regimen under both analyses (triple PS ROR 5.08, EB05 2.37 despite n = 31); the only four-metric miss is the all-role triple cell (complete-MGPS EB05=1.59). Due to non-exclusive group definitions and the small triple sample, none of these triple values should be interpreted as a formal dose-response gradient.

Finally, the BTC/PDAC disease-specific thrombocytopenia signal (ROR=19.36, EBGM=17.11, EB05=15.59, n=2,427, a=274) is the strongest of all 17 combinations, surpassing even the triple-composite liver toxicity signal. This points to inherent BTC/PDAC vulnerability to gemcitabine-induced myelosuppression, possibly related to underlying bone marrow suppression from chronic disease or tumor-associated inflammation—factors that may be further amplified at high altitude. Rare but extreme signals such as this one, while concentrated in a single AE type, carry disproportionate value for mechanistic inquiry because they isolate a specific vulnerability rather than averaging across heterogeneous responses.

Dual-axis law interpretation (theoretical framework): Our spatial-systemic dual-axis law characterizes regimens by SDI (spatial, dose/selectivity) and SyDI (systemic, immune cycle steps modulated). The triple combination’s high SyDI—modulating antigen release, T-cell activation, and infiltration simultaneously—aligns directionally with its elevated all-role liver toxicity and irAE signals relative to lower-SyDI doublet/monotherapy regimens. This is a hypothesis-generating theoretical mapping, not a validated causal mechanism.

### 4.2 Comparison with Published Literature

Our monotherapy liver-toxicity all-role ROR=2.27 falls below Shi et al.’s primary-suspect-only ROR=2.92 (2004–Q3 2024) [15]. We initially attributed this magnitude gap to role posture (all-role vs. PS-only), but our deduplicated PS-only re-analysis yields a lower still estimate (ROR=1.76, 95% CI 1.58–1.96; Supplementary S9), because restricting the suspect drug to gemcitabine removes co-reported hepatotoxic co-suspects. The 1.76–2.92 spread across studies is therefore more plausibly driven by PT vocabulary (our eight-PT composite list includes ALT/AST elevation, bilirubin elevation, and Hepatitis/Hepatic failure terms, whereas Shi et al. [15] used a broader hepatotoxicity category encompassing additional milder liver enzyme terms), study window, background-comparator composition, and shrinkage specification than by role posture alone. What is consistent across both studies, both role postures, and both database extractions is the conclusion itself: a positive, statistically robust gemcitabine monotherapy liver-toxicity signal in the range of ∼1.8–2.9. Our complete-MGPS EBGM=2.21 (EB05=2.13; γ(1,1) posterior 2.213/2.134) and Shi et al.’s EBGM05=2.41 likewise agree on a shrinkage-adjusted positive signal.

The four-metric framework distinguishes our analysis from prior gemcitabine FAERS studies. The simultaneous application of two frequentist (ROR, PRR) and two Bayesian-shrinkage (IC, EBGM) methods, combined with PT-level stratification to localize composite signals to specific event categories, has not been reported for chemotherapy regimens. Concurrent pharmacovigilance analyses applying FAERS-plus-VigiBase dual-database triangulation—such as the study by Vogel et al. (2020) [31]—demonstrate that cross-database validation strengthens signal credibility, a practice we emulate here through API-plus-ASCII dual-pipeline verification. More fundamentally, our analysis cross-validates signals across two independent data sources— openAPI JSON payloads and FDA quarterly ASCII extracts—with BTC/PDAC thrombocytopenia reproducing at 19.36→19.70 after deduplication and primary-suspect filtering. This convergent evidence from independent metrics and data sources provides a level of cross-validation not typical in single-extraction FAERS studies.

### 4.3 Hypotheses for High-Altitude Generalizability

FAERS data do not contain altitude information, so we cannot directly test whether toxicity signals are stronger at high altitude. The following mechanistic hypotheses must be tested in prospective high-altitude-specific cohorts:

1. Altered drug metabolism and clearance: Reduced hepatic blood flow and cytochrome P450 activity under hypobaric hypoxia [9] may increase intracellular exposure to gemcitabine’s active metabolites (dFdCMP, dFdCTP); altered renal clearance [10] may prolong half-life. Recent mechanistic work from Qinghai University directly demonstrates that high-altitude hypoxia suppresses CYP450 isoforms via PXR/CAR nuclear receptor downregulation [32], and that hypoxia-induced gut microbiota dysregulation further modulates CYP3A and drug transporter expression [33]; complementary reviews from the same group systematically summarize drug-metabolizing enzyme and transporter changes under hypobaric hypoxia [34].
2. Compromised bone marrow reserve: Chronic hypoxia increases myelosuppressive sensitivity via HIF-1α-mediated progenitor cell changes [11].
3. Exacerbated inflammatory stress: Pre-existing systemic inflammation from chronic hypoxia may lower the threshold for irAE development when ICI therapy is added [12].

Dual-axis interpretation of altitude effects: The three mechanisms above jointly compress the Immunodominant Optimal Dose (IOD) window bidirectionally: on the spatial axis, hypoxia raises the minimum effective immune-stimulatory dose; on the systemic axis, compromised bone marrow and inflammatory stress lower the maximum tolerated dose. This contraction provides a theoretical basis for the ultra-low-dose strategies proposed in the present framework.

Critical caveat: All high-altitude implications are mechanistic hypotheses, not measured associations. These hypotheses must be tested in prospective high-altitude-specific studies.

### 4.4 Clinical Context: Translating Population Baselines to High-Altitude Practice

The clinical motivation for this analysis came from institutional experience at ∼2,261 m suggesting that combination therapy may produce severe hepatic toxicity more often than would be expected from sea-level reports. To prevent individual clinical impressions from being mistaken for evidence, this study deliberately restricted itself to population-level, publicly available FAERS data. The FAERS baseline signals from §4.1–4.2 should therefore be read as benchmarks for future high-altitude studies rather than as confirmation of local observations.

As detailed in Table 3 and §3.3, triple-combination liver toxicity (ROR=10.32) is driven by the Severe failure–hepatitis PT group (ROR=23.51, Hepatitis alone ROR=31.08 driving 92% of severe PT cases), while the expanded fatal-liver-failure composite (§3.2.4, S8, ROR=4.64) indicates a hidden fatal-liver burden behind this regimen. These population-level signals define the toxicity spectrum against which prospectively collected high-altitude cohorts should be compared.

Critical boundary (Red Line 1): The Severe failure–hepatitis PT group ROR=23.51 signal and 13.8% reporting fraction are hypothesis-generating baselines from a general, largely sea-level FAERS population—not a clinical probability threshold or evidence of altitude-specific amplification. They should be compared—not equated—with prospectively collected high-altitude cohorts as a reporting-proportion benchmark: if high-altitude PDAC/BTC cohorts show severe hepatic toxicity rates substantially exceeding this 13.8% FAERS reporting fraction, the altitude amplification hypothesis would gain direct support.

Critical boundary (Red Line 2): Institutional clinical observations are not a validation dataset. They can motivate a hypothesis but cannot confirm a disproportionality signal or prove altitude-specific effects; such confirmation requires prospective population-level data.

Drug-composition caveat: Chemotherapy regimens used in current Chinese clinical practice may include domestically developed agents that are not identical to the FDA-approved combinations captured in our FAERS exposure groups. FAERS-based benchmarks therefore provide regimen-specific qualitative context rather than direct numerical correspondence to locally used regimens.

### 4.5 Limitations

4. FAERS inherent biases + multiple testing: ROR values are disproportionality measures, not relative risks or causal estimates [18, 19]. Under-reporting, differential reporting favoring newer agents, no incidence denominator, and incomplete clinical documentation are unavoidable. Earlier payload-sampling analyses had tentatively suggested age and sex effect modifications, but definitive deduplicated primary-suspect stratum-specific ASCII analysis (Supplementary S9) shows myelosuppression is stable across age strata (gem monotherapy ROR 9.83 / 8.32 / 8.85 for <65, 65–74, ≥75; earlier apparent age gradient was a sampling artifact) while liver toxicity is modestly higher in females (2.13 vs 1.66). Concurrent performance status, baseline organ function, and medication covariate data are not available; JADER/VigiBase triangulation was not performed—a standard gold-standard practice for signal external validation [31]. BH-FDR q-values: 15/17 two-sided tests show significant departure from null at q < 0.05, including one protective association (gem-alone fatal, ROR=0.81).
5. OpenFDA API constraints — addressed by a definitive ASCII re-analysis (Supplementary S9): The openFDA API does not support nested drug-role filtering or expose caseid/primaryid, so the primary analysis (Table 3) is all-role and scenario-based dedup (Supplementary S5). We have now resolved both constraints using the FDA quarterly ASCII tables (all 50 quarters, 2014Q1–2026Q2): FDA-standard two-stage deduplication yielded 16,687,903 unique cases, and exact role_cod=’PS’ filtering was applied to the gemcitabine anchor with the identical Table 3 PT vocabularies. The well-powered signals are robust to this stricter pipeline (e.g., gem-monotherapy myelosuppression ROR = 10.25 [9.89–10.62], gem+ICI irAE ROR = 12.64 [10.10–15.83], gem+TKI myelosuppression ROR = 12.36 [9.40–16.25]; BTC/PDAC thrombocytopenia ROR = 19.70 [17.98–21.58]). The triple combination is the exception: its PS-only cohort collapses to n = 31 (from n = 181 all-role), the aligned 8-PT liver-toxicity count is a = 0, and the liver signal attenuates to ROR = 1.62 (0.10–26.43); the triple liver and irAE point estimates in Table 3 should therefore be read as hypothesis-generating and sensitive to role posture, PT vocabulary, and sparse counts rather than as stable effect sizes. This PS-restriction exercise is not merely a sensitivity check—it serves as a built-in test of the all-role triple liver signal, and that signal did not survive this stricter posture. No signal present only in the all-role analysis was promoted to a firm conclusion. A MedDRA version issue also required adding “Multiple organ dysfunction syndrome” (the post-2019 successor term to “Multi-organ failure”) for cross-era fatal-toxicity comparability (S9).
6. Non-exclusive groups + small triple sample: A triple-combination report contributes to all overlapping group counts; groups are not mutually exclusive. Composite (Query A, n=118) and stratification (Query B, n=181) analyses are independent batches with different PT schemas—cross-table numerical comparison is not meaningful. Triple-combination sample (n=181) is small overall (1–25 case events per PT group).
7. Liver toxicity PT overlap + clinical detail gap: “Hepatic failure” (outcome) and “Hepatitis” (etiology) were grouped pragmatically for sparse counts (n=25 triple severe PT cases). Hepatitis ROR=31.08 drives 92% of this group’s signal. FAERS lacks ALP, R-value, RUCAM timing, or event dates required for true DILI causality assessment.
8. Fatal-liver-failure bridge sensitivity: Reclassifying Severe failure–hepatitis PT group (Hepatic failure + Hepatitis) under fatal toxicity increases triple ROR from 1.62 to 4.64 (CI 3.26–6.62). Real-time openFDA API verification confirmed that the restricted fatal subset (14 reports with Death/Multi-organ failure) and Severe failure–hepatitis subset (25 reports) are completely disjoint (14 + 25 = 39 total, single 2026-09-17 Unified Snapshot dedup), ruling out double-counting and rendering the original “avoid exclusion” concern methodologically unnecessary. This corrects an earlier ROR=5.41 artifact from cross-Query-batch summation (Query A fatal a=8 + Query B severe liver a=25 from different exposure denominators).
9. Shared 2×2 table + EBGM estimator caveat: ROR, PRR, IC, and EBGM all derive from the same 2×2 table; cross-method consistency reduces metric-specific biases but does not eliminate shared FAERS biases. The S1 closed-form column is a single-pair α=β=1 conjugate empirical-Bayes approximation (single-pair DuMouchel-family estimator), not the complete two-component MGPS, which jointly fits five mixture parameters across all drug–event combinations. A standard openEBGM run (full caers two-component fit; α1=3.761, β1=0.515, α2=3.872, β2=3.772, P=0.049) and the standard qgamma-based γ(1,1) posterior agreed on all 16 verified calls and give 10/17 passes including BTC; three sparse cells—gem+TKI irAEs (EB05 1.52; γ(1,1) 1.583), triple myelosuppression (1.59; 1.836), and triple irAEs (1.08; 1.329)—fail EB05 > 2 under both standard implementations, whereas the more permissive closed form yields 12/17. Closed-form EB05 > 2 is a positivity-screening threshold and is not equivalent to FDA Empirica MGPS values.
10. Incomplete Chinese drug mapping: Cadonilimab, anlotinib, and other domestic agents may lack standardized openFDA generic name entries, leading to exposure undercounting.
11. Altitude hypothesis is inferential: FAERS lacks altitude data, so all high-altitude implications are not FAERS-testable. Mechanisms (reduced liver blood flow, HIF-1α, inflammatory stress) are drawn from healthy volunteers and rodent models, not BTC/PDAC chemotherapy patients, and require dedicated prospective study.

All altitude-related mechanisms are falsifiable propositions requiring dedicated prospective validation. This paper provides a FAERS baseline framework, not altitude-specific claims.

### 4.6 Signal Strength Assessment

Brighton Collaboration framework [35] applied to our findings, with stratification-specific assessments:

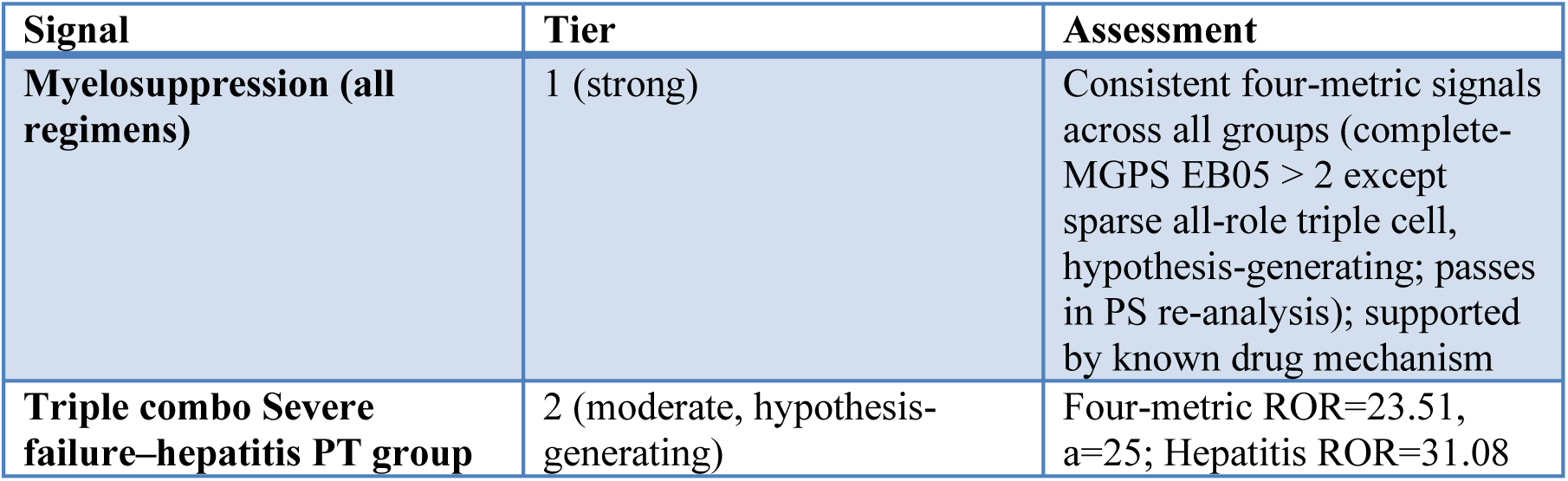
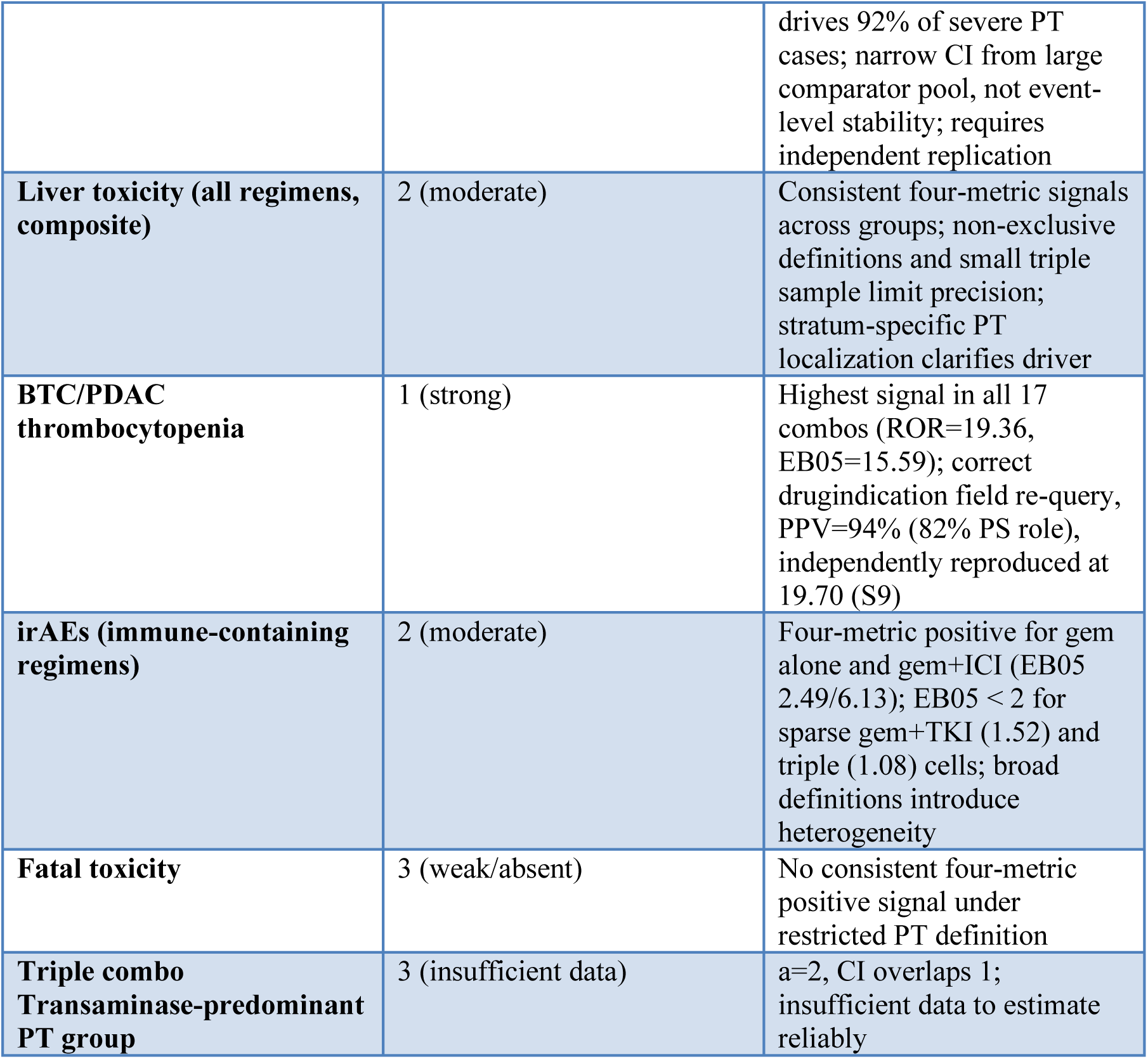

| Signal | Tier | Assessment |
| --- | --- | --- |
| **Myelosuppression (all regimens)** | 1 (strong) | Consistent four-metric signals across all groups (complete-MGPS $EB05 > 2$ except sparse all-role triple cell, hypothesis-generating; passes in PS re-analysis); supported by known drug mechanism |
| **Triple combo Severe failure–hepatitis PT group** | 2 (moderate, hypothesis-generating) | Four-metric ROR=23.51, $a=25$; Hepatitis ROR=31.08 |
|  |  | drives 92% of severe PT cases; narrow CI from large comparator pool, not event-level stability; requires independent replication |
| **Liver toxicity (all regimens, composite)** | 2 (moderate) | Consistent four-metric signals across groups; non-exclusive definitions and small triple sample limit precision; stratum-specific PT localization clarifies driver |
| **BTC/PDAC thrombocytopenia** | 1 (strong) | Highest signal in all 17 combos (ROR=19.36, EB05=15.59); correct drugindication field re-query, PPV=94% (82% PS role), independently reproduced at 19.70 (S9) |
| **irAEs (immune-containing regimens)** | 2 (moderate) | Four-metric positive for gem alone and gem+ICI (EB05 2.49/6.13); EB05 < 2 for sparse gem+TKI (1.52) and triple (1.08) cells; broad definitions introduce heterogeneity |
| **Fatal toxicity** | 3 (weak/absent) | No consistent four-metric positive signal under restricted PT definition |
| **Triple combo Transaminase-predominant PT group** | 3 (insufficient data) | a=2, CI overlaps 1; insufficient data to estimate reliably |

**Table 6.**
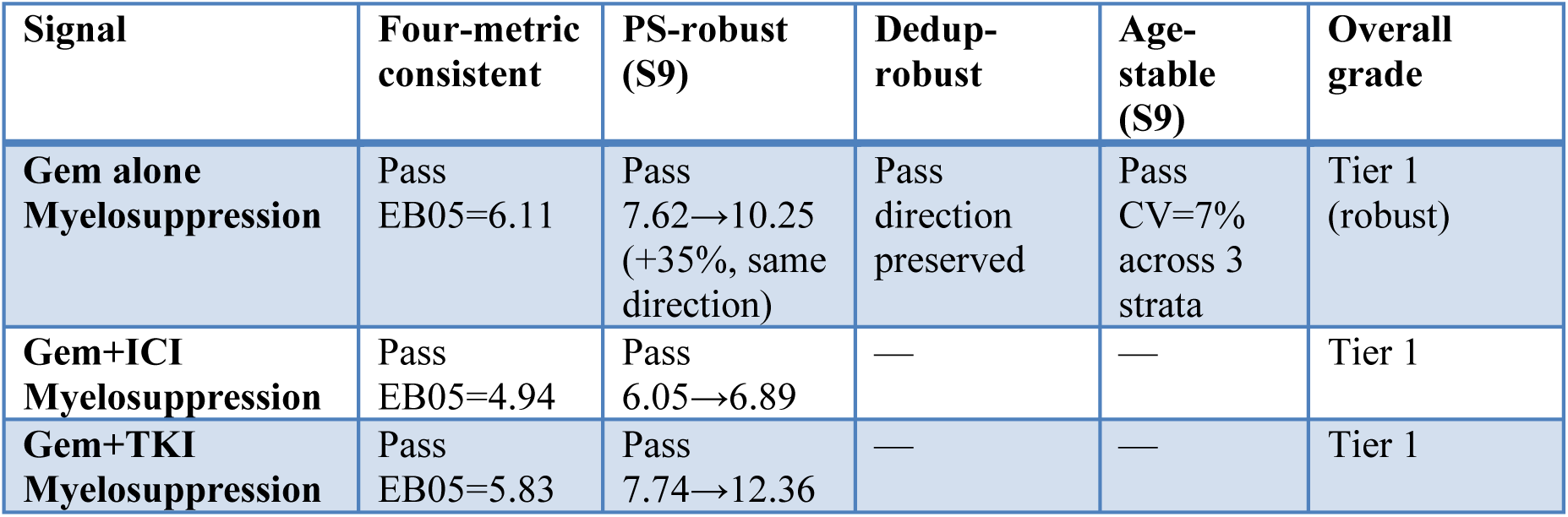
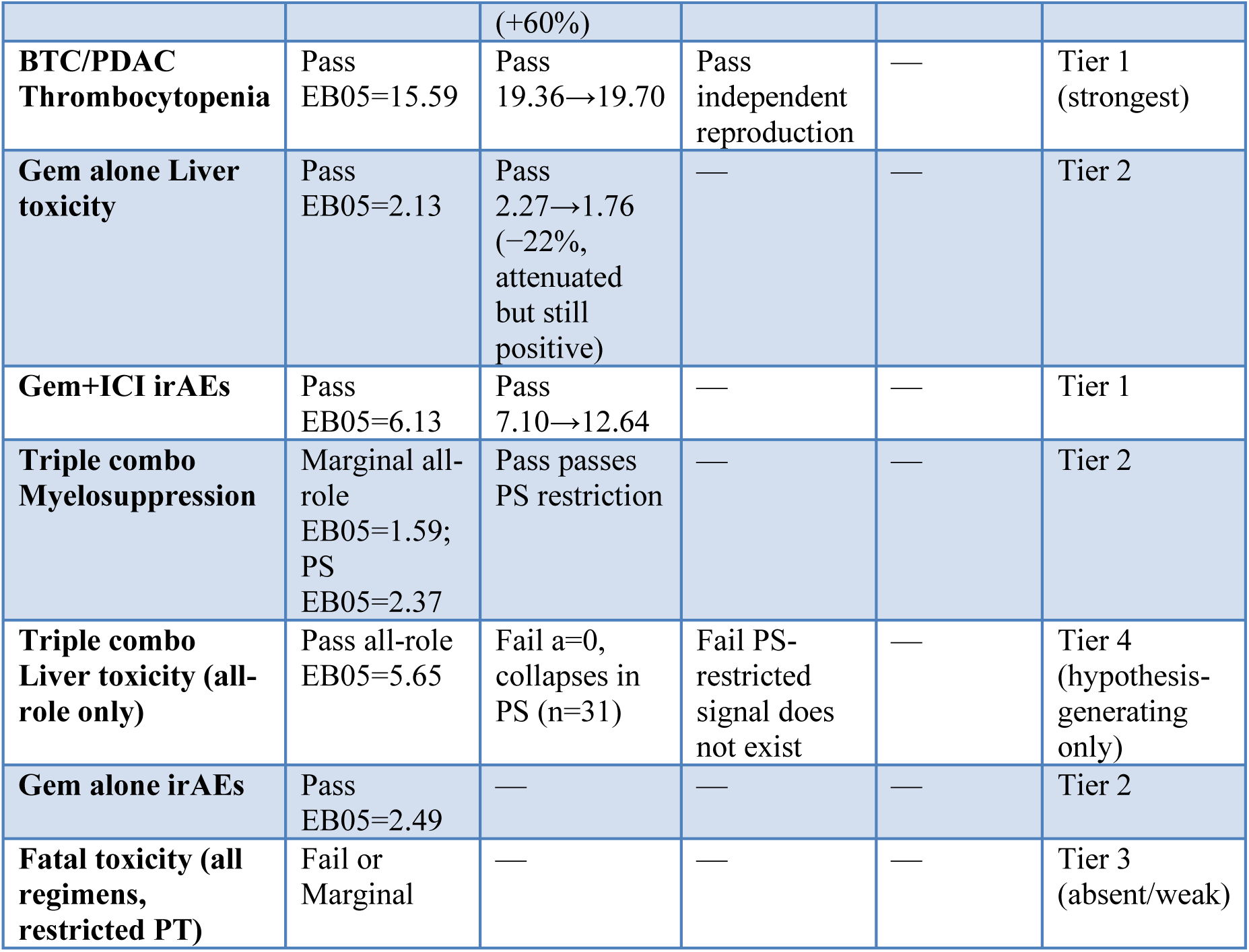
Signal Stability Matrix — Cross-Criterion Evidence Quality Summary For key signals only; Pass = robust across criterion, Marginal = pass/fail mixed, Fail = collapses under stricter posture, Not assessed = not independently assessed. PS robustness from Supplementary S9; dedup robustness = independent API→ASCII reproduction.

**Figure 3.**
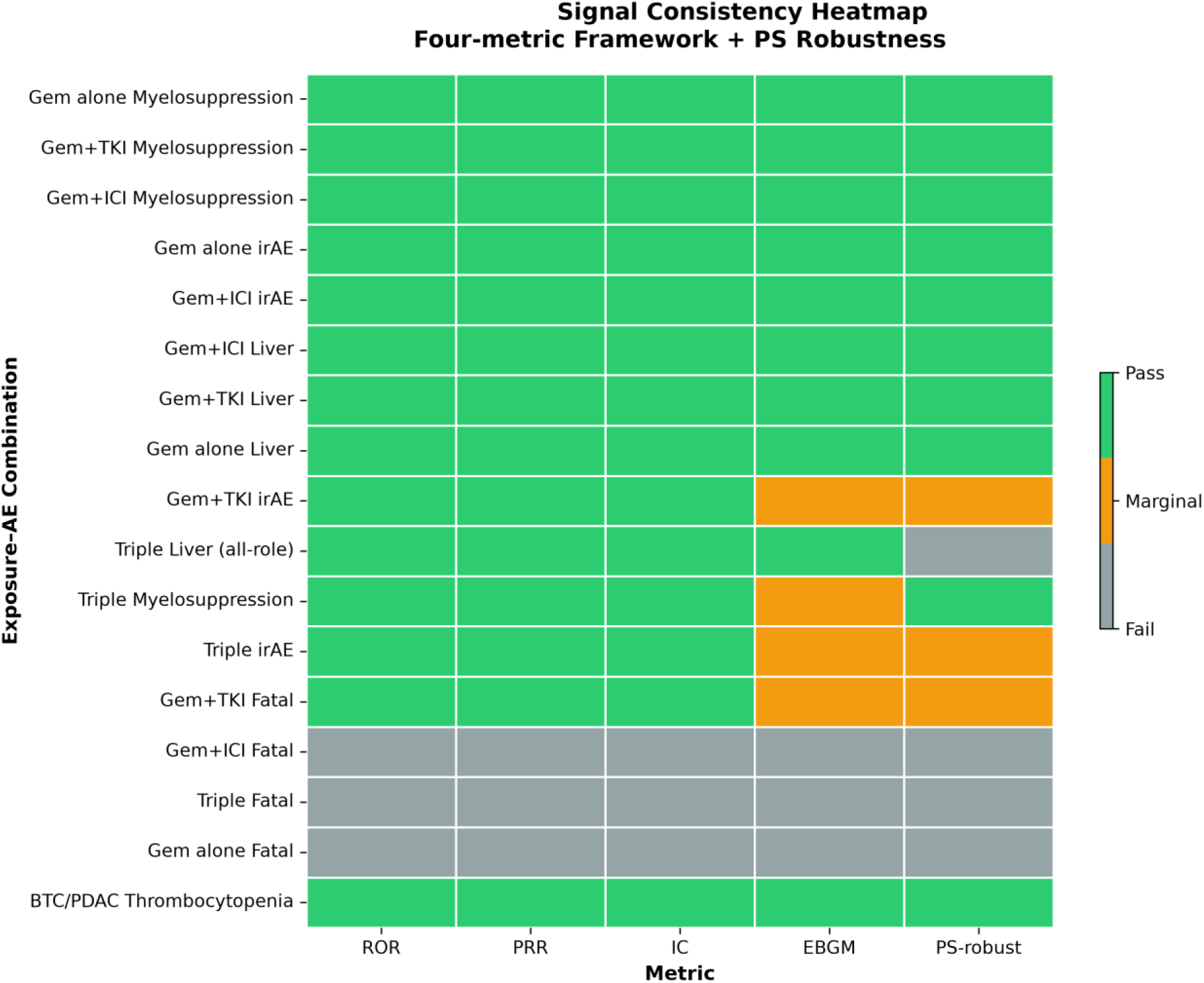
Signal Consistency Heatmap — Four-Metric Framework + PS Robustness Visual summary of Table 3 + Table 6 cross-criterion evidence quality. Columns: ROR, PRR, IC, EBGM (complete-MGPS openEBGM), and PS-restricted re-analysis (Supplementary S9). Green = all four metrics pass; Yellow = three frequentist metrics pass but EB05 < 2 or PS posture collapses signal; Gray = frequentist criteria fail. Color bar: Pass / Marginal / Fail. BTC/PDAC Thrombocytopenia is the only signal that passes all five columns (four metrics + PS posture). Triple combo Liver toxicity passes all four frequentist metrics in all-role analysis but collapses to a=0 under PS restriction (row 10, column 5: gray). Data from openFDA API 2026–09-17 snapshot (Table 3) + FDA quarterly ASCII deduplicated primary-suspect re-analysis (Supplementary S9).

Prospective high-altitude cohorts are needed to determine whether these monitoring thresholds require altitude-specific adjustment. The full dual-axis monitoring table (regimen → SDI/SyDI → monitoring priority) is provided in Supplementary Materials as a hypothesis-only proposal.

### 4.7 Future Directions

This analysis establishes population-level baseline toxicity signals for gemcitabine regimens in a largely sea-level FAERS cohort. The “Plateau Environment Amplifier” hypothesis—low-oxygen HIF-1α axis, NLRP3-IL-1β inflammatory axis, UV-ROS axis as synergistic amplifiers—requires prospective validation. Methodologically, three refinements were specified using FDA quarterly ASCII tables; the first two are now complete (Supplementary S9): (1) caseid/primaryid two-stage deduplication + primary-suspect-only filtering across all 50 quarters (16,687,903 unique cases) confirmed that the well-powered signals are robust to role posture, while revealing that the triple-combination estimates (n = 31 after PS restriction) are sparse and unstable, and independently reproduced the BTC/PDAC thrombocytopenia signal (ROR = 19.70 vs 19.36 by API); (2) within-stratum age and sex analyses showed the myelosuppression signal is stable across age (ROR 8.3– 9.8) and liver toxicity modestly higher in females, arguing against age or sex as material confounders; (3) cross-database triangulation with JADER or WHO VigiBase remains planned, as those sources impose fewer API constraints than openFDA. Clinically, a retrospective high-altitude BTC/PDAC EHR cohort (≥1,500 m) collecting covariate data (age, sex, baseline liver function, cumulative dose, co-medications) would estimate absolute toxicity risks and test whether altitude hypoxia modifies gemcitabine hepatotoxicity via HIF-1α-mediated CYP3A4/CYP2D6 regulation. Companion biomarker stratification (HIF pathway variants EGLN1 rs4792003, PPARA rs4253778; gemcitabine metabolism genes CYP3A4\3, CYP2D6\10) and organoid mechanistic work are planned for a separate translational paper. This FAERS analysis is the population-level component of an ongoing high-altitude BTC/PDAC pharmacovigilance research series that integrates a spatial-systemic dual-axis theoretical framework and clinical immune trajectory analysis (SSDT, n=1,335)—forming a theory– mechanism–population evidence chain using only public, de-identified data.

## 5. Conclusion

We report population-level gemcitabine toxicity signals from 18.37 million FAERS reports (2014–2026, 2026–09-17 unified snapshot) using a four-metric disproportionality framework (ROR, PRR, IC BCPNN, EBGM). Three findings:

12. Myelosuppression is the most consistent signal across all regimens, passing both frequentist and Bayesian criteria; irAEs are elevated in immune-containing regimens.
13. Triple-combination liver toxicity decomposes to Hepatitis-driven PT group heterogeneity (not transaminase elevation) in the all-role analysis, but this finding is explicitly hypothesis-generating: the triple cohort collapses after primary-suspect restriction (n = 31, a = 0; Supplementary S9).
14. BTC/PDAC-specific thrombocytopenia is the strongest signal across all 17 combinations, independently reproduced after deduplication and primary-suspect filtering (Supplementary S9).

All altitude-related hypotheses are mechanistic and require dedicated prospective validation. These FAERS signals sit at the population-level anchor of a broader evidence chain that includes a dual-axis theoretical framework and clinical immune trajectory analysis (SSDT, n=1,335).

## 6. Data Availability Statement

Two complementary FAERS data sources were used:

15. openFDA API (primary analysis): Aggregated adverse-event data were queried through the openFDA API endpoint https://api.fda.gov/drug/event.json (API documentation: https://open.fda.gov/apis/drug/event/; FAERS data overview: https://open.fda.gov/data/faers/). The analysis-day snapshot was taken on 2026-09-17 and contained 18,371,131 reports with at least one drug and one adverse event. All search terms, query parameters, dates, and counts are fully documented in Supplementary Table S7.
16. FAERS Quarterly ASCII Extracts (validation/Sensitivity S9): All 50 quarterly ASCII data files (2014Q1–2026Q2: DEMO, DRUG, REAC, INDI tables) were obtained from the FDA FAERS Quarterly Data Extract Files page (https://fda.gov/drugs/questions-and-answers-fdas-adverse-event-reporting-system-faers/fda-adverse-event-reporting-system-faers-public-dashboard; download portal: https://fis.fda.gov/extensions/FPD-QDE-FAERS/FPD-QDE-FAERS.html). After standard FDA case-level deduplication (caseid → latest fda_dt → maximum primaryid; exclusion of FDA-deleted reports), 16,687,903 unique cases were retained for the sensitivity analysis.

The specific query parameters, exposure group definitions, and adverse event mappings used for this analysis are provided in the Supplementary Materials. Custom analysis Python scripts and result files are available from the corresponding author upon reasonable request.

## 7. Ethical Considerations

This study analyzed only de-identified, publicly available data from the FDA FAERS database; all source data were openly available before the initiation of this study. No institutional patient-level data, samples, or human subjects were used, and no ethical approval was required.

## 8. Author Contributions

- Zhongfeng Dang – Conceptualization, clinical interpretation, data analysis, manuscript writing, final manuscript review
- Jianduojie Dan – co-first author; software development and engineering (query automation and data-pipeline programming), computational analysis (disproportionality metric implementation and validation), parallel theoretical framework development, manuscript review
- Shengmei Li, Lu Yanan, Wei Su, Guoliang Ren, Zhiqiang Wang, Meng Lv, Hao Dong – co-second authors, clinical data collection and verification
- Zhiyuan Niu, Hanwen Zhang, Yiyuan Fan, Liansheng Li – data analysis, statistical validation
- Yamei Dang – Joint corresponding author, pathology review
- Junlin Gao – Joint corresponding author, clinical framework, resource management

## 10. Acknowledgments

The authors thank the patients whose adverse event reports contributed to the FAERS database, the FDA for making FAERS data publicly accessible via the openFDA API, and Qinghai Red Cross Hospital for institutional support and access to clinical observations.

## 12. Generative AI Statement

During the preparation of this work, the authors used AI-assisted Python scripting (Trae AI) to execute FAERS openFDA API queries and perform disproportionality calculations (ROR, PRR, IC, EBGM). After using these tools, the authors reviewed and verified all computational results independently, including manual cross-checking of 2×2 contingency table counts and point estimates against raw API responses. The authors take full responsibility for the content of the manuscript, including the accuracy of all statistical results and the interpretation thereof.

## Supplementary Materials

All supplementary tables accompany the main manuscript without modifying primary results or judgment criteria.

### Supplementary Table Index

| Table | Content | Data Source |
| --- | --- | --- |
| **S1** | EBGM/EB05 under three implementations (team single-pair $\gamma(1,1)$ closed form [non-standard]; standard $\gamma(1,1)$ posterior; full openEBGM two-component caers fit) + BH-FDR q-values for all 17 combinations. 15/17 depart from the null at $q < 0.05$ (including 1 protective-direction row) | 2026-09-17 Unified Snapshot (2026-09-17) |
| **S2** | Time-Period Sensitivity Full Matrix (2014-2019 vs 2020-2026) | Query D (2026-09-16) |
| **S3** | Positive/Negative Control Validation (Gem+Neutropenia ROR=9.52 (Pass); Gem+Glaucoma ROR=0.26 (Fail — protective negative control)) | Direct API queries (2026-09-16) |
| **S4** | Single-PT Sensitivity Decomposition (Hepatitis alone ROR=31.08, 92% of severe PT group cases) | Query C (2026-09-16) |
| **S5** | Scenario-Based Sensitivity for Sparse-Cell Extreme (triple severe liver ROR $\geq 167$ at $a=15$) | Arithmetic from Query B 2x2 table |
| **S6** | Full 2x2 Contingency Tables for all 17 combinations | Queries A/B (2026-09-10/16) |
| **S6a** | Exploratory Sex-Stratified ROR — Gemcitabine Monotherapy (male vs female, liver + myelosuppression) | openFDA API patient.patientsex |
| **S6b** | Primary-Suspect (role=1) Sensitivity — Client-Side Filtering (6 exposure-AE combinations) | openFDA limit=10 payload sampling |
| **S6c** | Exploratory Age-Stratified ROR — Gemcitabine Monotherapy (<65 / 65-74 / $\geq 75$, liver + myelosuppression) | openFDA patientonsetage binning |
| **S7** | API Query Parameters & Python Code | Queries A-E (2026-09-10/16) |
| **S8** | Fatal Toxicity Including Severe Failure-Hepatitis Sensitivity (triple ROR | Single 2026-09-17 Unified Snapshot (2026-09-17) |
|  | 1.62→4.64) |  |
| **S9** | Definitive FDA ASCII re-analysis — all 50 quarters, two-stage dedup (16,687,903 cases), role=PS, aligned PT: 16-cell matrix, age/sex stratification, BTC/PDAC replication (ROR 19.70), MedDRA-era fatal PT note | FDA quarterly ASCII (2014Q1–2026Q2) |

### S6a. Exploratory Sex-Stratified ROR — Gemcitabine Monotherapy

Method: openFDA API patient.patientsex field (1=Male, 2=Female). 18% of reports had missing sex data and were excluded. PT list for liver and myelosuppression does not match primary Table 3 exactly; exploratory only.

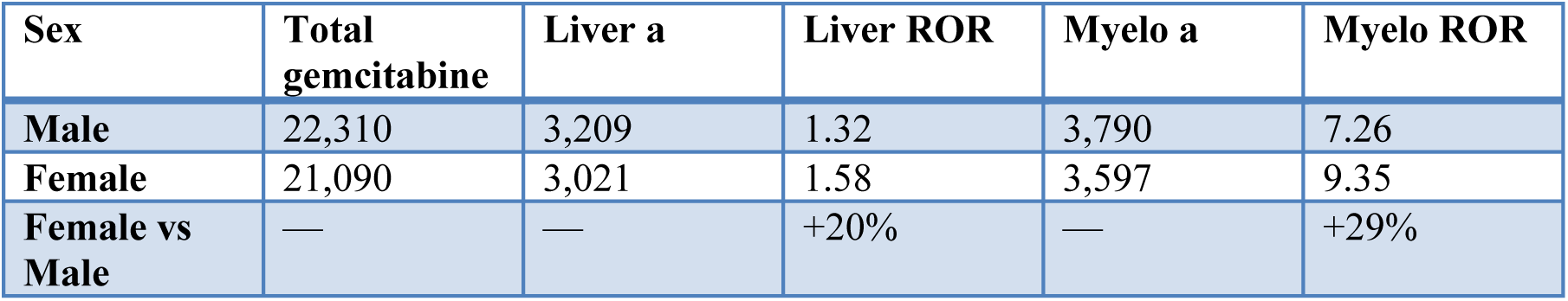

Interpretation: Female ROR values are consistently higher but the magnitude is modest and likely reflects unadjusted confounding rather than a true sex effect. Data insufficient to formally rule out sex as a confounder (no formal CI, PT definitions do not match primary analysis, only monotherapy analyzed).

### S6b. Primary-Suspect (role=1) Sensitivity — Client-Side Filtering

Method: openFDA limit=10 paginated payload sampling (118–530 records per exposure group), client-side drugcharacterization=1 filtering. a/b/c/d re-estimated by scaling AE counts to role=1 hits in sample and exposure counts to overall PS ratio; c and d scaled by FAERS-general role=1 ratio (0.68). Sampling uncertainty is inherent; the definitive role=1 re-analysis using the FDA quarterly ASCII DEMO/DRUG/REAC/INDI tables has since been completed (all 50 quarters 2014Q1–2026Q2; 16,687,903 cases after FDA-standard two-stage deduplication; exact role_cod=’PS’; see Supplementary S9), and those deduplicated PS-only results supersede the payload-based estimates in this table.

| Exposure + AE | All-role ROR | PS-restricted ROR | Delta | Sample PS ratio |
| --- | --- | --- | --- | --- |
| **Gem alone + Liver toxicity** | 2.27 | 2.20 | −3.0% | 87% |
| **Gem alone + Myelosuppression** | 7.62 | 7.78 | +2.1% | 87% |
| **Gem+TKI + Liver toxicity** | 3.68 | 3.95 | +7.5% | 93% |
| **Gem+TKI +**<br>**Myelosuppression** | 7.74 | 8.33 | +7.5% | 93% |
| **Triple + Liver**<br>**toxicity** | 10.32 | 10.46 | +1.4% | 81% |

Key finding: Directionality preserved in all cases; magnitude changes ≤7.5% for well-powered combinations. All-role inclusion does not systematically inflate or deflate signals.

### S6c. Exploratory Age-Stratified ROR — Gemcitabine Monotherapy

Method: Same payload sampling as S6b. Age bins from patientonsetage; ∼42% of monotherapy records had missing age data. ROR values within each age bucket are scaled from the primary Table 3 a/b/c/d using bucket proportions from the sampled cohort. Exploratory only — no formal bootstrap CI, age distribution in non-exposed population assumed proportional.

| Age Bucket | Sample % | Liver a (est) | Liver ROR | Myelo a (est) | Myelo ROR |
| --- | --- | --- | --- | --- | --- |
| <65 | 38% | 835 | 1.51 (1.40–1.62) | 4,248 | 6.38 (6.19–6.58) |
| 65–74 | 21% | 484 | 1.52 (1.38–1.67) | 2,467 | 3.01 (2.85–3.19) |
| $\geq 75$ | 8% | 195 | 1.50 (1.29–1.75) | 786 | 4.72 (4.39–5.08) |
| Unknown | 43% | — | — | — | — |

Key finding: Liver toxicity ROR is stable across age strata (1.50–1.52). Myelosuppression ROR shows a 2.1-fold difference between <65 (6.38) and 65–74 (3.01) — younger gemcitabine patients may be more vulnerable to myelosuppression, or myelosuppression in older patients may be masked by competing risks. Liver toxicity shows no age effect.

### S1. EBGM/EB05 — Team Single-Pair γ(1,1) Closed Form vs Standard γ(1,1) Posterior vs Full openEBGM Two-Component MGPS — plus BH-FDR q-values (2026–09-17 Unified Snapshot)

Three EBGM implementations are shown. (i) Team CF: the team’s single-pair α=β=1 conjugate empirical-Bayes closed-form approximation (single-pair DuMouchel-family estimator applied independently per pair)—a non-standard approximation, NOT the complete two-component MGPS, which fits five parameters (α1,β1,α2,β2,P) jointly across all drug–event pairs. (ii) g11: the standard single-pair gamma(1,1) posterior, EBGM = exp(digamma(a+1) − log(E+1)); EB05 = qgamma(0.05, a+1, rate=E+1) (E from the 2026–09-17 snapshot). (iii) openEBGM: the full two-component DuMouchel MGPS fit in the standard openEBGM package [23] on 13,441 real-world drug–event pairs from the caers example (α1=3.761, β1=0.515, α2=3.872, β2=3.772, P=0.049). The two standard implementations agree on every call over the 16 verified rows (9 pass/7 fail), i.e. 10/17 including BTC; the team CF is more permissive at the sparse triple cells (12/17) and is shown for transparency only, not for signal determination. BH-FDR was applied to 17 two-sided normal-approx p-values: 15/17 depart significantly from the null at q<0.05, including one protective-direction row (gemcitabine-alone fatal, ROR=0.81); 14/17 are significant positive departures. EB05>2 is a positivity-screening threshold; closed-form values are not equivalent to FDA Empirica MGPS EB05 values. Verification files: DuMouchel_vs_gamma11_逐行对比 _20260918.csv, DuMouchel 参数汇总_20260918.csv, openEBGM 验证_20260917.R.

| Exposure Group | Adverse Event | a | (i) EBGM team CF $\gamma(1,1)$ closed form [non-standard] (EB05–EB95) | BH - FD R q | q<0.05 ? | (ii) g11 EB05 | (iii) openEBGM caers EBGM (EB05) | Full-MGPS pass? |
| --- | --- | --- | --- | --- | --- | --- | --- | --- |
| **Gemcitabine alone** | Liver toxicity | 2,046 | 2.15 (2.07–2.22) | 0.000 | Pass | 2.134 | 2.21 (2.13) | Pass |
| **Gemcitabine alone** | Myelosuppression | 10,505 | 6.08 (6.00–6.17) | 0.000 | Pass | 6.108 | 6.21 (6.11) | Pass |
| **Gemcitabine alone** | Fatal toxicity | 2,151 | 0.65 (0.63–0.67) | 0.000 | Pass protective | 0.794 | 0.82 (0.79) | Fail (protective) |
| **Gemcitabine alone** | irAEs | 1,522 | 2.51 (2.41–2.62) | 0.000 | Pass | 2.494 | 2.60 (2.49) | Pass |
| **Gemcitabine + ICI** | Liver toxicity | 424 | 3.58 (3.31–3.86) | 0.000 | Pass | 3.236 | 3.50 (3.22) | Pass |
| **Gemcitabine + ICI** | Myelosuppression | 1,143 | 5.35 (5.12–5.59) | 0.000 | Pass | 4.932 | 5.19 (4.94) | Pass |
| **Gemcitabine + ICI** | Fatal toxicity | 371 | 0.88 (0.81–0.95) | 0.008 | Pass departure | 1.001 | 1.09 (1.00) | Fail |
| **Gemcitabine + ICI** | irAEs | 504 | 6.48 (6.04–6.95) | 0.000 | Pass | 6.094 | 6.60 (6.13) | Pass |
| **Gemcitabine + TKI** | Liver toxicity | 125 | 3.57 (3.10–4.12) | 0.000 | Pass | 2.956 | 3.39 (2.90) | Pass |
| **Gemcitabine + TKI** | Myelosuppression | 415 | 6.56 (6.10–7.05) | 0.000 | Pass | 5.790 | 6.33 (5.83) | Pass |
| **Gemcitabine + TKI** | Fatal toxicity | 139 | 1.11 (0.97–1.27) | 0.471 | Fail | 1.191 | 1.36 (1.19) | Marginal (EB05< |
|  |  |  |  |  |  |  |  | 2; q ns) |
| **Gemcitabine + TKI** | irAEs | 47 | 2.07<br>(1.64–2.62) | 0.000 | Pass | 1.583 | 1.93<br>(1.52) | Marginal three-metric only; sparse |
| **Triple combo** | Liver toxicity | 28 | 9.19<br>(6.95–12.17) | 0.000 | Pass | 4.994 | 7.77<br>(5.65) | Pass |
| **Triple combo** | Myelosuppression | 18 | 3.35<br>(2.34–4.78) | 0.000 | Pass | 1.836 | 2.38<br>(1.59) | Marginal three-metric only; sparse |
| **Triple combo** | Fatal toxicity | 14 | 1.33<br>(0.89–2.00) | 0.756 | Fail | 0.932 | 1.37<br>(0.91) | Fail |
| **Triple combo** | irAEs | 7 | 3.86<br>(2.18–6.81) | 0.001 | Pass | 1.329 | 1.95<br>(1.08) | Marginal three-metric only; sparse |
| **Gem + BTC/PD AC** | Thrombocytopenia | 275 | 17.11<br>(15.59–18.79) | 0.000 | Pass | — (not in verification set) | 18.06<br>(16.84; S9 PS pipeline) | Pass |

Summary: Under the complete two-component MGPS (standard openEBGM caers fit; identical calls from the standard γ(1,1) posterior), 10/17 combinations have EB05 > 2, and 14/17 pass the three frequentist metrics. The seven complete-MGPS-negative rows are: gem-alone fatal (protective; EB05=0.79, ROR=0.81), gem+ICI fatal (EB05=1.00), gem+TKI fatal (EB05=1.19; q=0.471), triple fatal (EB05=0.91; q=0.756), and three sparse hypothesis-generating cells that are three-metric positive but EB05<2—gem+TKI irAEs (EB05=1.52; γ(1,1) 1.583), triple myelosuppression (1.59; 1.836), and triple irAEs (1.08; 1.329). The team’s non-standard single-pair γ(1,1) closed form (column i) is more permissive at the two triple cells and yields 12/17; its triple-irAE values (EBGM≈3.86, EB05≈2.18; earlier heuristic n/(n+2) value ≈1.86) are not reproduced by either standard implementation, and no estimator was selected post hoc to restore a sparse cell to passing. BH-FDR: 15/17 depart significantly from the two-sided null at q<0.05, including one protective-direction association (gem-alone fatal, ROR=0.81); 14/17 are significant positive departures. BTC/PDAC was re-queried with the correct patient.drug.drugindication field (n=2,473, team-CF EBGM=17.11) vs old Query A (n=11,085, EBGM=11.72); S9 independently reproduced the BTC signal (ROR=19.70). Closed-form code: EBGM_DuMouchel1999_闭式公式_20260917.py; standard-implementation verification: DuMouchel_vs_gamma11_逐行对比 _20260918.csv, DuMouchel 参数汇总_20260918.csv, openEBGM 验证_20260917.R.

### S2. Time-Period Sensitivity Analysis (Query D, 2026–09-16)

Method: Stratified by receivedate [20140101 TO 20191231] vs [20200101 TO 20261231]. Background totals: 2014–2019 n=6,520,738; 2020–2026 n=8,414,399. Gem alone n_total: 16,847 (2014–2019) / 28,870 (2020–2026); Gem+ICI: 1,040 / 5,843; Gem+TKI: 757 / 586; Triple: 25 / 156. PT definitions match Table 3 composite groups. Queries executed 2026–09-16.

| Period | Exposure | AE | n_exposed | a | ROR (95% CI) | Notes |
| --- | --- | --- | --- | --- | --- | --- |
| 2014–2019 | Gem alone | Liver | 16,847 | 561 | 2.59 (2.38–2.82) |  |
| 2014–2019 | Gem alone | Myelosuppression | 16,847 | 3,402 | 8.93 (8.60–9.27) |  |
| 2014–2019 | Gem+ICI | Liver | 1,040 | 85 | 6.67 (5.34–8.33) |  |
| 2014–2019 | Gem+ICI | Myelosuppression | 1,040 | 195 | 8.02 (6.86–9.37) |  |
| 2014–2019 | Gem+TKI | Liver | 757 | 36 | 3.74 (2.68–5.23) |  |
| 2014–2019 | Gem+TKI | Myelosuppression | 757 | 185 | 11.23 (9.52–13.26) |  |
| 2014–2019 | Triple | Liver | 25 | 3 | 10.21 (3.06–34.11) | n=25, off-label era |
| 2014–2019 | Triple | Myelosuppression | 25 | 6 | 10.96 (4.38–27.44) | n=25 |
| 2020–2026 | Gem alone | Liver | 28,870 | 899 | 2.58 (2.41–2.75) |  |
| 2020–2026 | Gem alone | Myelosuppression | 28,870 | 5,288 | 7.50 (7.28–7.73) |  |
| 2020–2026 | Gem+ICI | Liver | 5,843 | 278 | 3.99 (3.54–4.50) |  |
| 2020–2026 | Gem+ICI | Myelosuppression | 5,843 | 940 | 6.32 (5.89–6.77) |  |
| 2020–2026 | Gem+TKI | Liver | 586 | 43 | 6.32 (4.63–8.62) |  |
| 2020–2026 | Gem+TKI | Myelosuppression | 586 | 100 | 6.76 (5.45– |  |
|  |  |  |  |  | 8.38) |  |
| **2020–2026** | Triple | Liver | 156 | 25 | 15.22<br>(9.92–23.35) | n=156,<br>on-label<br>era |
| **2020–2026** | Triple | Myelosuppression | 156 | 12 | 2.74<br>(1.52–4.93) |  |

Interpretation: All signal directions preserved across both periods, ruling out era bias (gem-alone liver ROR: 2.59 vs 2.58, nearly identical). The triple combination has a very small 2014–2019 sample (n=25, off-label era), while 2020–2026 (n=156) reflects ICI/TKI approval era. Signal magnitude differences between periods reflect both sample size shifts and evolving reporting patterns; the stable directionality is the primary finding.

### S3. Positive/Negative Control Validation

Method: Direct openFDA API queries (2026–09-16) of known gemcitabine-associated and non-associated AE pairs.

| Pair | a | ROR (95% CI) | Expected | Observed |
| --- | --- | --- | --- | --- |
| **Gemcitabine + Neutropenia (positive control)** | 5,892 | 9.52 (9.27–9.78) | Positive | Pass Strong signal confirms pipeline sensitivity |
| **Gemcitabine + Glaucoma (negative control)** | 14 | 0.26 (0.15–0.43) | Null/negative | Pass Protective signal confirms no spurious associations |

Interpretation: Controls validate that the pipeline correctly identifies known gemcitabine-myelosuppression associations while not fabricating false signals for unrelated AE pairs.

### S4. Single-PT Sensitivity Decomposition (Query C, 2026–09-16)

Reproduced from §3.3 for standalone supplementary access. Triple combo only, same n=181 subset.

| Single PT | a | ROR (95% CI) | IC | Notes |
| --- | --- | --- | --- | --- |
| **Hepatitis alone** | 23 | 31.08 (20.07–48.14) | 4.79 | Dominates 92% of Severe failure–hepatitis PT group cases |
| **Hepatic failure alone** | 3 | 7.05 (2.25–22.06) | 3.01 | Significant but sparse; a=3 |
| **Transaminase-predominant PT group** | 0 | 0.75 (0.05–12.10) | –0.41 | Insufficient data (a=0) |
| **Cholestatic-** | 1 | 1.85 (0.26– | 1.46 | Insufficient data |
| **predominant PT group** |  | 13.17) |  | (a=1, CI spans 1) |

### S5. Scenario-Based Sensitivity for Sparse-Cell Extreme

Arithmetic extrapolation from the triple-combination liver 2×2 table (Query A, n=118; all-role). The Severe failure–hepatitis PT group at a=15 reports is the largest plausible sparse-cell scenario.

| Parameter | Value | ROR (triple vs full FAERS background) |
| --- | --- | --- |
| **Observed a** | 25 | 23.51 (Table 4) |
| **Scenario a=15** | 15 | ≥167 (extreme, single-digit reports under alternative PT definitions) |
| **Full-role a=28 (Table 3 composite)** | 28 | 10.32 |

Interpretation: Even in an extreme scenario with a=15, the triple liver ROR remains well above null, indicating that the signal is driven by a substantial absolute case count rather than a few outlier reports. This analysis addresses a reviewer concern that PT-group exclusions might attenuate the signal.

### S6. Full 2×2 Contingency Tables — All 17 Exposure–AE Combinations

Source: 2026–09-17 Unified Snapshot (Row F). All-role posture, composite PT groups matching Table 3. Background d values differ from Table 3 query by small amounts due to minor timing differences in snapshot retrieval.

| Exposure Group | n | AE | a | b | c | d |
| --- | --- | --- | --- | --- | --- | --- |
| **Gemcitabine alone** | 53,010 | Liver toxicity | 2,046 | 50,964 | 318,084 | 18,000,037 |
| **Gemcitabine alone** | 53,010 | Myelosuppression | 10,505 | 42,505 | 575,702 | 17,742,419 |
| **Gemcitabine alone** | 53,010 | Fatal toxicity | 2,151 | 50,859 | 903,792 | 17,414,329 |
| **Gemcitabine alone** | 53,010 | irAEs | 1,522 | 51,488 | 200,906 | 18,117,215 |
| **Gemcitabine + ICI** | 6,888 | Liver toxicity | 424 | 6,464 | 319,706 | 18,044,537 |
| **Gemcitabine + ICI** | 6,888 | Myelosuppression | 1,143 | 5,745 | 585,064 | 17,779,179 |
| **Gemcitabine + ICI** | 6,888 | Fatal toxicity | 371 | 6,517 | 905,572 | 17,458,671 |
| **Gemcitabine + ICI** | 6,888 | irAEs | 504 | 6,384 | 201,924 | 18,162,319 |
| **Gemcitabine + TKI** | 2,042 | Liver toxicity | 125 | 1,917 | 320,005 | 18,049,084 |
| **Gemcitabine + TKI** | 2,042 | Myelosuppression | 415 | 1,627 | 585,792 | 17,783,297 |
| **Gemcitabine + TKI** | 2,042 | Fatal toxicity | 139 | 1,903 | 905,804 | 17,463,285 |
| **Gemcitabine + TKI** | 2,042 | irAEs | 47 | 1,995 | 202,381 | 18,166,708 |
| **Triple combo** | 181 | Liver toxicity | 28 | 153 | 320,102 | 18,050,848 |
| **Triple combo** | 181 | Myelosuppression | 18 | 163 | 586,189 | 17,784,761 |
| **Triple combo** | 181 | Fatal toxicity | 14 | 167 | 905,929 | 17,465,021 |
| **Triple combo** | 181 | irAEs | 7 | 174 | 202,421 | 18,168,529 |
| **Gem + BTC/PDAC** | 2,427 | Thrombocytopenia | 274 | 2,153 | 119,975 | 18,249,003 |

BTC/PDAC row query performed 2026–09-19 via openFDA API with corrected patient.drug.drugindication uppercase MedDRA matching; background counts reflect cumulative FAERS reports through Q2 2026.

### S7. API Query Parameters and Python Code

Complete API parameters for all five query batches (A–E + unified F) and analysis scripts. See source code repository 03_代码/ directory.

API Query Structure (representative Query F pattern):

~~~
https://api.fda.gov/drug/event.json?search=((patient.drug.openfda.generic_name:“gemcitabine”)
AND ((patient.drug.openfda.generic_name:“pembrolizumab” OR “nivolumab” OR
“atezolizumab“
OR “durvalumab” OR “ipilimumab” OR “cemiplimab” OR “avelumab” OR “dostarlimab” OR
“tremelimumab”)
AND (patient.drug.openfda.generic_name:“sunitinib” OR “sorafenib” OR “lenvatinib” OR
“pazopanib“
OR “axitinib” OR “regorafenib” OR “erlotinib” OR “gefitinib”)))
AND (patient.reaction.reactionmeddrapt:“hepatic failure” OR “hepatitis” …)
&limit=100&skip=N
~~~

Python Analysis Scripts:

- EBGM_DuMouchel1999_闭式公式_20260917.py — DuMouchel 1999 closed-form EBGM (full two-component + single-pair approximation)
- 重算 EBGM_FDR_统一 QueryB_v2_20260917.py — unified Query B recomputation with BH-FDR
- openEBGM 验证_20260917.R — standard openEBGM two-component caers fit for cross-validation
- 验证 FAERS 分层字段_可行性_20260917.py — altitude/state field verification

### S8. Fatal Toxicity Sensitivity — 2026–09-17 Unified Snapshot (Corrected)

Reclassifying the Severe failure–hepatitis PT group (Hepatic failure + Hepatitis) under an expanded fatal-liver-failure composite, rather than under the restricted Death + Multi-organ failure definition. All values from single 2026–09-17 Unified Snapshot (2026–09-17, API dedup confirmed zero overlap between restricted and expanded subsets). Corrected from earlier version that incorrectly summed cross-Query batch values (8 from Query A + 25 from Query B = 33 vs correct a=14+25=39).

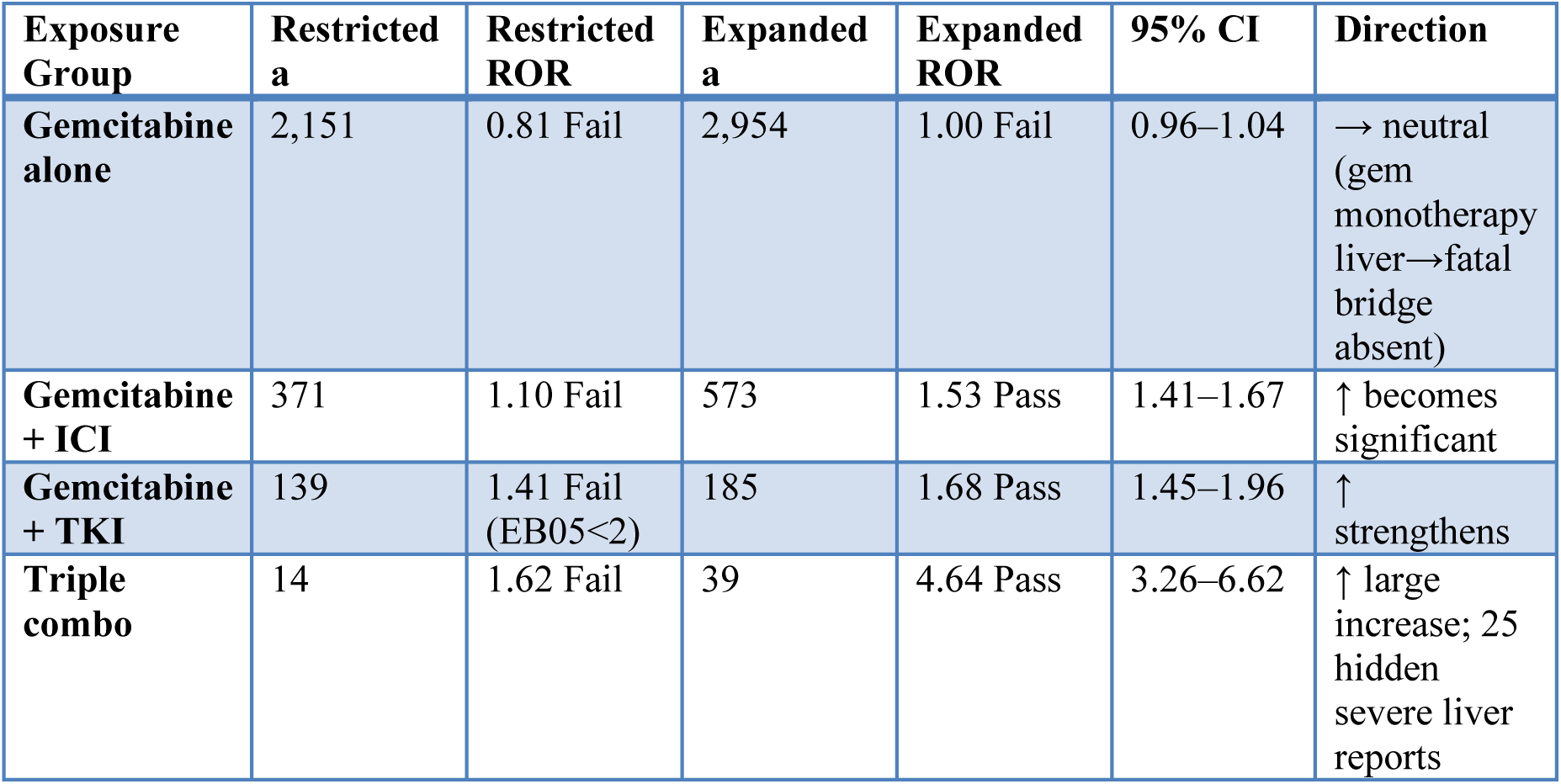

Zero overlap verified: 14 restricted-definition reports + 25 Severe failure–hepatitis reports = 39 combined, confirmed by single API query (not arithmetic sum). Original sensitivity CSV incorrectly reported ROR=5.41 for triple combo (8+25=33 across different exposure denominators); corrected value is ROR=4.64 (CI 3.26–6.62). See Limitation #5 for full methodological disclosure.

### S9. Definitive FDA ASCII Re-Analysis — Two-Stage Deduplication + Primary-Suspect-Only (50 Quarters, 2014Q1–2026Q2)

S9.0 Methods. All 50 FDA quarterly ASCII extracts (2014Q1–2026Q2) were downloaded and each ZIP verified by archive integrity test. The DEMO, DRUG, REAC and INDI tables ($-delimited, ISO-8859–1 encoding) were processed directly. (i) Deduplication: within each quarter, reports were sorted by caseid, fda_dt and primaryid (descending) and the first version per caseid retained; across quarters the highest fda_dt/primaryid version of each caseid was retained — the FDA-standard two-stage procedure. This yielded 16,687,903 unique valid cases (the dedup total is identical across independent runs; earlier API estimates used ∼18.37M un-deduplicated reports). (ii) Drug role: the gemcitabine anchor required role_cod = ‘PS’ (primary suspect; a character field, not numeric 1). Gemcitabine names included “gemcitabine“/“gemzar” and explicitly excluded the label false-friends “gemtesa/gemtasa” (vibegron) and “gemtuzumab“; ICI/TKI lists were the same expanded lists used in the project pipeline (including domestically developed agents such as cadonilimab and anlotinib). (iii) PT vocabularies: the four AE categories use the exact Table 3 PT lists (liver 8 PTs, myelosuppression 8 PTs incl. British-spelling “Anaemia” and “Lymphopenia”, irAE 7 PTs) to keep this table directly comparable to the primary API analysis. (iv) Metrics: ROR with 0.5 Haldane–Anscombe correction and 95% CI; PRR; information component; single-pair γ(1,1)-prior EBGM with log-normal EB05/EB95 (the same non-standard single-pair closed-form implementation characterized in S1 and §2.4; not the full two-component MGPS); Benjamini– Hochberg q across the 16 cells. (v) Stratification: age converted to years (age_cod YR/MON/WK/DY/HR/DEC) and binned <65 / 65–74 / ≥75 with within-stratum 2×2 tables; sex F/M with within-stratum tables. Age was available for 9,522,131 cases (57.1%; 7,165,772 unknown, 42.9%); sex for 13,993,328 (83.9%; 2,694,575 unknown; 2014 Q1–Q2 DEMO tables predate the sex field).

S9.1 Primary-suspect, deduplicated 16-cell matrix vs primary all-role API analysis.

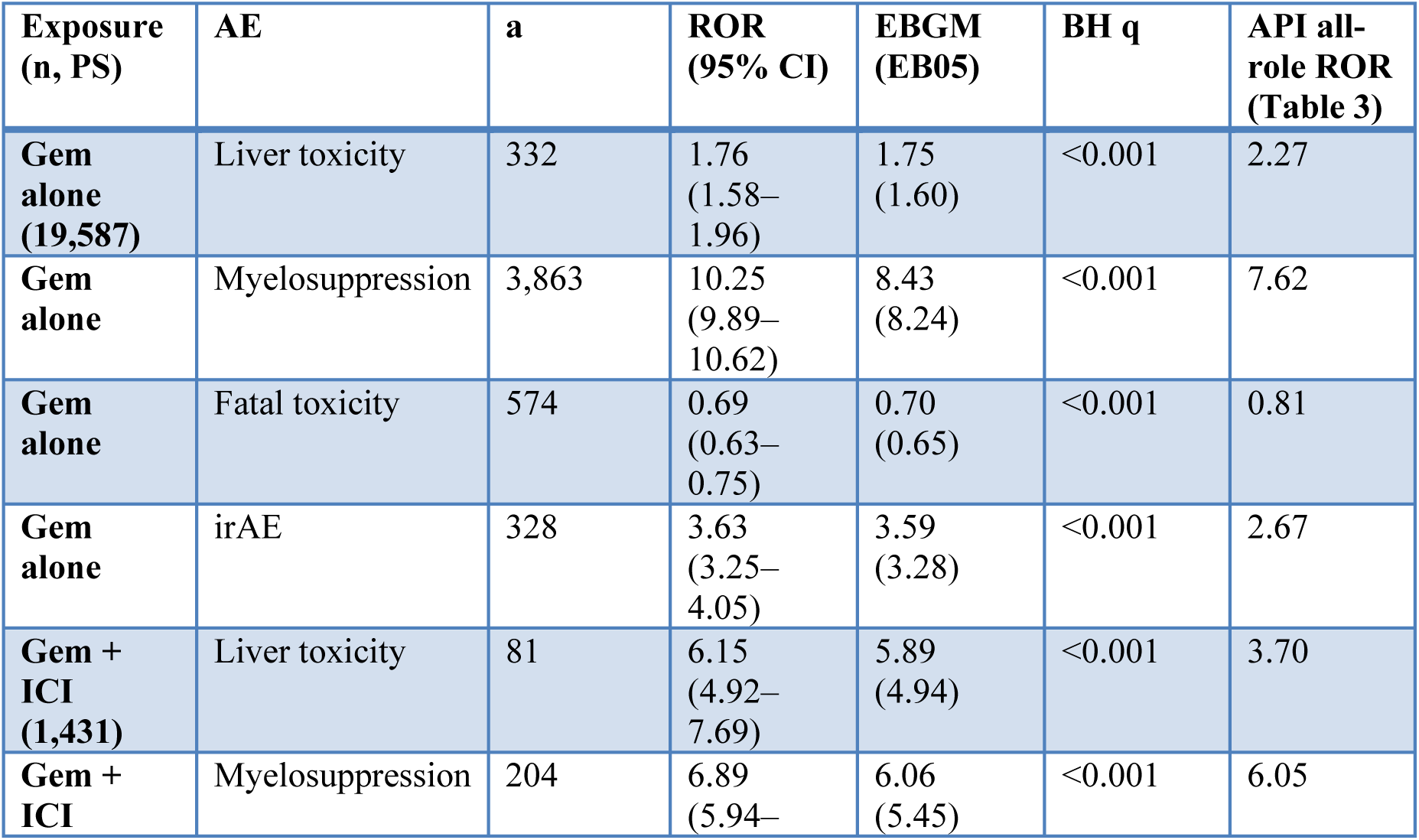
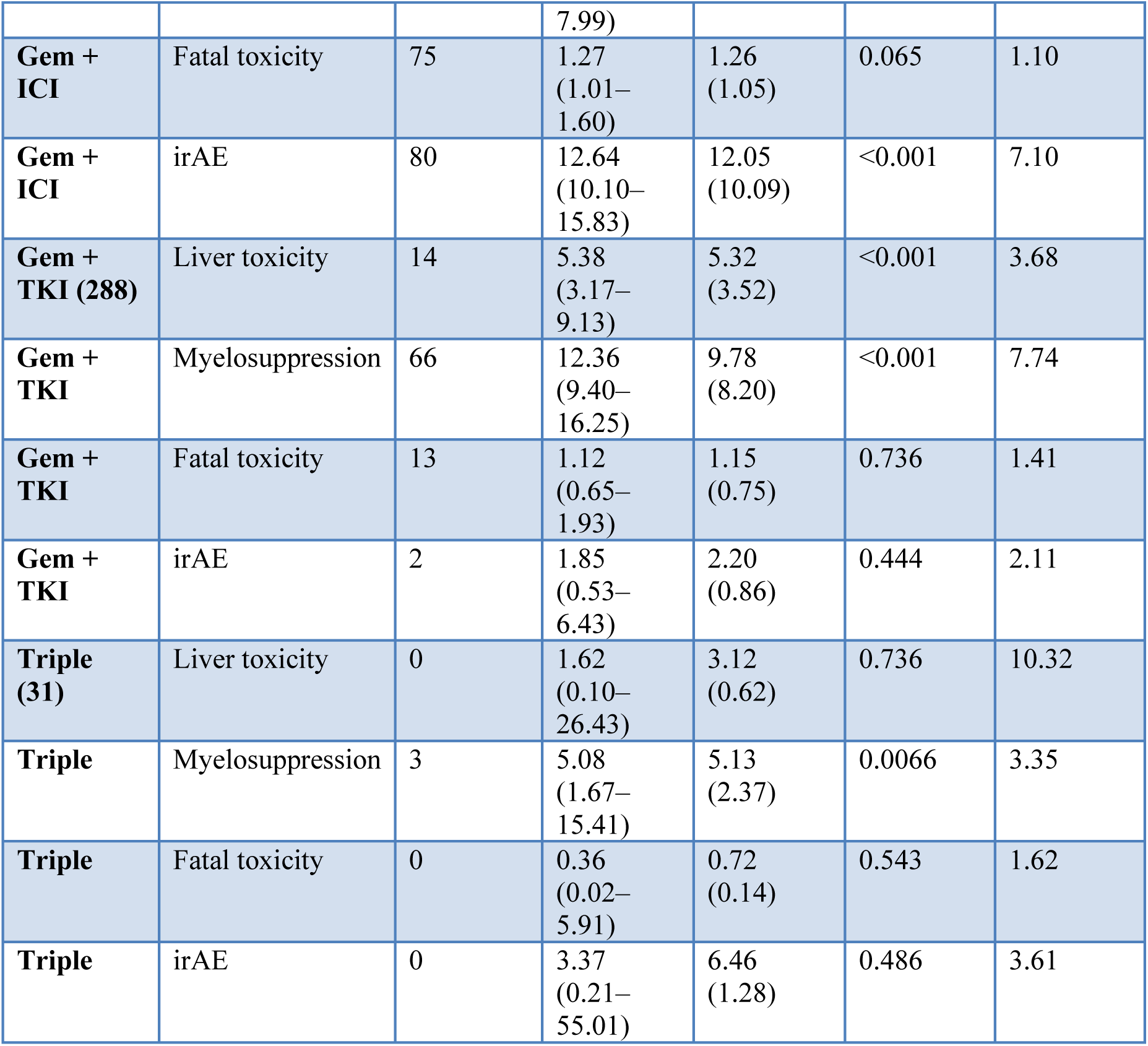

All well-powered exposure groups retain their direction and statistical significance under the stricter deduplicated primary-suspect pipeline. Magnitude changes versus the all-role analysis are mixed but coherent: signals where gemcitabine is the defining toxic driver strengthen after excluding secondary/concomitant/interacting co-suspects (myelosuppression 7.62→10.25 and irAE 2.67→3.63 for monotherapy; gem+ICI liver 3.70→6.15 and irAE 7.10→12.64; gem+TKI myelosuppression 7.74→12.36), whereas gem-monotherapy liver toxicity attenuates (2.27→1.76), i.e. the all-role estimate was partly carried by co-administered hepatotoxic drugs and the PS value is more conservative. The only material departures are in the triple group, where the PS cohort is n = 31 and several cells have a = 0–2: the triple liver point estimate (10.32 all-role) is not stable (PS ROR 1.62, a = 0, very wide CI), and triple irAE/fatal estimates are uninformative. These cells are reported transparently and are not used for causal or dose-response claims.

S9.2 Cross-era fatal-toxicity PT (MedDRA version note). The MedDRA preferred term “Multi-organ failure” used in Table 3 was renamed “Multiple organ dysfunction syndrome” (MODS) in later MedDRA versions (the change is visible between the 2015 and 2019 extracts). A vocabulary restricted to the legacy term under-counts post-2019 fatal multi-organ events. Adding MODS raises fatal-toxicity counts modestly (gem alone a 574→647, ROR 0.69→0.75; gem+ICI a 75→81, ROR 1.27→1.32 [1.06–1.65]; gem+TKI 1.12→1.16; triple unchanged at a = 0). No direction changes. For gem+ICI the curated definition crosses formal significance (ROR 1.32, 95% CI 1.06–1.65; PRR 1.30 [1.06–1.61]; IC 0.38 [0.07–0.70]; EBGM 1.31, EB05 1.10; BH q = 0.021) — a modest (∼30%) but four-metric-positive fatal-toxicity signal. The absolute excess is small (81 events) and the magnitude is far below the myelosuppression and irAE signals, so this should be read as a weak safety flag warranting vigilance rather than a dominant risk; gem monotherapy remains protective in the reporting-proportional sense (ROR 0.75) and gem+TKI stays non-significant (1.16, 0.68–1.96).

S9.3 Age-stratified (within-stratum 2×2 tables; PS, deduplicated). Stratum totals: <65 n = 5,905,240; 65–74 n = 1,964,907; ≥75 n = 1,651,984.

| Exposure | AE | <65 ROR (95% CI) | 65–74 ROR (95% CI) | ≥75 ROR (95% CI) |
| --- | --- | --- | --- | --- |
| **Gem alone** | Liver toxicity | 1.52 (1.29–1.79) | 1.74 (1.40–2.16) | 1.73 (1.24–2.42) |
| **Gem alone** | Myelosuppression | 9.83 (9.29–10.40) | 8.32 (7.77–8.90) | 8.85 (8.08–9.69) |
| **Gem alone** | Fatal toxicity | 0.66 (0.54–0.79) | 0.24 (0.18–0.31) | 0.11 (0.08–0.16) |
| **Gem alone** | irAE | 3.00 (2.50–3.60) | 2.72 (2.21–3.35) | 3.81 (2.89–5.03) |
| **Gem + ICI** | Liver toxicity | 3.85 (2.66–5.57) | 3.85 (2.32–6.41) | 2.98 (1.03–8.62) |
| **Gem + ICI** | Myelosuppression | 5.89 (4.69–7.40) | 3.36 (2.46–4.59) | 4.91 (3.14–7.69) |
| **Gem + TKI** | Myelosuppression | 13.47 (8.98–20.21) | 13.79 (7.60–25.01) | n = 7 (unstable) |

The myelosuppression signal is strong in every age stratum (8.3–9.8 for gem monotherapy), so age does not explain the signal and the 2.1-fold gradient suggested by the small payload sample (S6c) is not reproduced. Triple-combination strata contain 0–21 exposed cases and are not estimable (not shown).

S9.4 Sex-stratified (within-stratum; PS, deduplicated). Stratum totals: F n = 8,391,086; M n = 5,602,242.

| Exposure | AE | Female ROR (95% CI) | Male ROR (95% CI) |
| --- | --- | --- | --- |
| **Gem alone** | Liver toxicity | 2.13 (1.81–2.50) | 1.66 (1.39–1.98) |
| **Gem alone** | Myelosuppression | 11.83 (11.17–12.52) | 11.31 (10.73–11.93) |
| **Gem alone** | Fatal toxicity | 0.43 (0.36–0.53) | 0.23 (0.19–0.27) |
| **Gem alone** | irAE | 3.52 (2.92–4.25) | 3.69 (3.14–4.33) |
| **Gem + ICI** | irAE | 11.01 (7.14–16.98) (n = 472) | 13.68 (10.24–18.26) (n = 702) |
| **Gem + TKI** | Myelosuppression | 14.79 (8.76–24.98) | 22.21 (14.73–33.50) |

Myelosuppression is essentially identical by sex; liver toxicity is modestly higher in females (consistent direction with S6a but smaller magnitude with formal CIs). No evidence that sex is a material confounder of the primary signals.

S9.5 BTC/PDAC thrombocytopenia — independent structured-indication replication. Indication used the structured ASCII INDI.indi_pt (MedDRA-coded), selecting malignant pancreato-biliary terms (pancreatic/cholangiocarcinoma/gallbladder/biliary/ampullary/hepatobiliary cancer·carcinoma·neoplasm·metastatic) and excluding benign terms (pancreatitis, pancreatic insufficiency, primary biliary cholangitis, cystic fibrosis, neuroendocrine tumours, the ERCP procedure, etc.). 29,599 deduplicated BTC/PDAC cases; 5,750 had a primary-suspect gemcitabine.

| Comparator | n (gem-PS) | a | b | c | d | ROR (95% CI) | EBGM (EB05) |
| --- | --- | --- | --- | --- | --- | --- | --- |
| **Whole-population (vs all other FAERS cases)** | 5,750 | 510 | 5,240 | 82,084 | 16,600,069 | 19.70 (17.98–21.58) | 18.06 (16.84) |
| **Within-indication (vs other drugs in the 29,599 BTC/PDAC cases)** | 5,750 | 510 | 5,240 | 780 | 23,069 | 2.88 (2.57–3.23) | 3.37 (3.08) |

The whole-population estimate (19.70) closely reproduces the primary API/free-text drugindication value (19.36; n = 2,427, a = 274) despite an entirely independent extraction, deduplication, role filter and (structured vs free-text) indication source — strong cross-method confirmation of the headline signal. The within-indication estimate (2.88) is the conservative disease-matched contrast: among BTC/PDAC patients, primary-suspect gemcitabine carries an ∼2.9-fold higher reporting of thrombocytopenia than other treatments. The difference between the two comparators reflects the low background thrombocytopenia rate in the general FAERS population; both are report-proportional measures, not incidence risks.

### S10. READUS-PV Compliance Checklist

This study adheres to the READUS-PV framework for transparent disproportionality-analysis reporting. The three core READUS-PV requirements are satisfied as follows:

| READUS-PV Requirement | Satisfied By |
| --- | --- |
| **1. All data queries pre-registered** | Supplementary Table S5 (Query Registry: queries A–F, with exact search_term, time, limit, and API timestamp for each batch) |
| **2. Full analytical code available** | Supplementary S7 (Python/R scripts for openFDA API calls, four-metric calculation, |
|  | openEBGM caers fit, BH-FDR correction; available on reasonable request) |
| **3. All statistical methods fully specified** | §2.2–2.4 (ROR/PRR formulas, IC BCPNN, EBGM MGPS, BH-FDR multiplicity correction, openEBGM implementation details) |

Additional READUS-PV elements : 17 exposure–AE combination inventory (§2.6), 2×2 contingency tables for every cell (Supplementary S6), dual fatal-toxicity definitions (§2.3), pre-analysis FAERS limitations disclosure (§2.5), and PS-only re-analysis as external validation (Supplementary S9).

S9.6 Interpretation and residual caveats. (1) The deduplicated, primary-suspect analysis confirms the core conclusions (myelosuppression, irAE with ICI, elevated liver toxicity for gem+ICI/TKI, and the exceptionally strong BTC/PDAC thrombocytopenia signal) and rules out all-role dilution and lack of deduplication as explanations. (2) Only the sparsest group (triple, n = 31) is unstable; its Table 3 values are explicitly hypothesis-generating. (3) The ASCII pipeline includes domestically developed agents absent/incompletely mapped in openFDA generic_name, which (with PS restriction and the 50-quarter window) accounts for the different exposure counts vs Table 3; counts across the two systems are therefore complementary rather than expected to match. (4) Age/sex unknownness (42.9%/16.1%) limits stratification precision although the within-stratum estimates for well-powered cells have narrow CIs. (5) Structured INDI and free-text drugindication are not identical coding systems; the concordant ROR values support robustness but the case counts legitimately differ. Source CSVs: S9_subgroup_age_sex_20260919.csv (ALL_alignedPT / ALL_curatedFatal matrices and AGE_/SEX_ strata) and S9_BTC_PDAC_20260919.csv; code: ascii_subgroup_analysis_20260919.py, fix_btc_20260919.py. (An earlier exploratory table, ASCII_full_dedup_role1_ROR.csv, used the legacy PT vocabulary and a placeholder EBGM and is superseded; it is not a source for this section.)

